# Identification of serum protein biomarkers in individuals with Niemann-Pick disease, type C1

**DOI:** 10.64898/2026.01.12.26343721

**Authors:** Khushboo Singhal, Matthew T. Menold, Niamh X. Cawley, Kiersten Campbell, Nicole Y. Farhat, Derek Alexander, Ryan K. Dale, Forbes D. Porter

**Affiliations:** Section on Molecular Dysmorphology, Division of Translational Research, Eunice Kennedy Shriver National Institute of Child Health and Human Development, National Institutes of Health, Bethesda, MD, USA; Bioinformatics and Scientific Programming Core, Eunice Kennedy Shriver National Institute of Child Health and Human Development, National Institutes of Health, Bethesda, MD, USA

**Keywords:** Niemann-Pick disease, type C1, NPC1, Neurodegeneration, Serum Protein Biomarkers, Proximal Extension Assay

## Abstract

**Background:** Niemann-Pick disease, type C1 (NPC1), is a rare, fatal, neurodegenerative lysosomal disorder caused by pathological variants in *NPC1*. Defects in lysosomal cholesterol transport result in the accumulation of unesterified cholesterol within the endo-lysosomal compartments. Delayed diagnosis, limited treatment options, and phenotypic heterogeneity characterized by a broad range of signs/symptoms underscore the urgent need for effective biomarkers to facilitate diagnosis, monitor disease progression and assess therapeutic response. The goal of this study was to identify serum protein biomarkers for NPC1.

**Methods:** Proximal Extension Assays (PEA) were used to determine relative protein expression levels from 68 serum samples from NPC1 individuals and 20 age-appropriate control serum samples. Statistical models identified NPC1 disease-specific effects after adjusting for covariates. Selected proteins were orthogonally validated by ELISA and correlated with assessments of both disease severity (Age of Neurological Onset (ANO) and Annual Severity Increment Score (ASIS)) and disease burden (NPC Neurological Severity Score (NSS).

**Results:** Quantifiable data was obtained on 2888 proteins, revealing 186 increased (adjusted log_2_FC ≥ 1) and 286 decreased (adjusted log_2_FC ≤ −1) proteins with adj. p-value < 0.1 when comparing NPC1 individuals not being treated with miglustat versus control serum samples. Using orthogonal assays, we confirmed significant elevations for seven proteins: TREM2, AgRP, CCL18, Cathepsin L, GPNMB, NPY, and HSD17B14, and a significant decrease of BDNF. We further identified 100 proteins whose abundance levels were significantly altered towards normal by miglustat treatment. We found the 17-domain NPC NSS to be correlated with protein levels in the PEA data. Orthogonally validated data correlated with the age of neurological onset. We also identified 25 differentially abundant serum proteins in NPC1 baseline samples which are predominantly expressed in brain regions.

**Conclusions:** The statistical analysis pipeline developed in this study is flexible and scalable and supports application to high-dimensional proteomic datasets. This study identified and validated serum proteins with altered expression in individuals with NPC1, responded to miglustat therapy, and correlated with disease severity or burden. These proteins may have clinical utility as biomarkers and provide insights into cellular mechanisms contributing to NPC1 disease pathology.

**Trial Registrations:** NCT00344331 (Registration on 2006-06-23)

## Background

Niemann-Pick disease, type C, is an ultra-rare, fatal, lysosomal storage disorder due to pathogenic variants in either *NPC1* (MIM 257220; ∼95% cases) or *NPC2* (MIM 607625; ∼5% cases)(1). The disease follows an autosomal recessive inheritance pattern, with an estimated incidence rate of about 1 in 100,000(2, 3). NPC1 is involved in intracellular cholesterol transport, and pathological variations in *NPC1* lead to impairment in the transport of cholesterol out of the endo-lysosomal compartment, resulting in luminal storage of unesterified cholesterol and glycosphingolipids(4). In addition to cellular stress caused by lysosomal dysfunction, the cellular bioavailability of cholesterol is also reduced. These two pathological processes have detrimental effects on cellular function that ultimately contribute to neurodegeneration(5).

Pathologically, NPC1 disease is characterized by progressive neurodegenerative loss of Purkinje neurons (contributing to cerebellar ataxia), synaptic disturbance, and myelination defects, among others(6). Phenotypically, NPC1 disease is highly heterogeneous, with variability in the age of neurological onset (ranging from prenatal stages to adulthood), disease severity, symptoms, and survival outcomes. Neurodegeneration typically progresses over time, with early neurological onsets indicating a more rapid decline. Depending on the age of the individual and disease severity, symptoms can be visceral (hepatosplenomegaly), neurological, cognitive, and psychiatric(6). This pronounced heterogeneity highlights the importance of considering these variables when identifying reliable biomarkers for disease progression and therapeutic response. Laboratory diagnosis of NPC1 is based on a combination of blood-based testing for biochemical biomarkers (oxysterols, lyso-sphingomyelin 509, and bile acid metabolites) and molecular testing(7). There is a significant diagnostic delay, and the initial presentation of the disease may be confused with that of other lysosomal storage disorders with similar presentations(6, 8).

The US Food and Drug Administration (FDA) recently approved arimoclomol (Myplyffa®) in combination with miglustat (Zavesca®) for treating NPC1(9). It has been proposed that arimoclomol improves lysosomal membrane stability by inducing heat shock proteins, thereby boosting the ability of these lysosomes to break down overabundant lipids and protect from cell death(9). Miglustat is a glycosphingolipid synthesis inhibitor that has long been approved for NPC1 treatment in European countries(10). In the feline model of NPC1, miglustat has been shown to increase Purkinje cell survival, modulate the microglial phenotype, and reduce glycosphingolipid accumulation(11). In individuals with NPC1, the benefits of miglustat have been reported for improving cellular function, stabilizing clinical phenotypes, stabilizing swallowing function, reducing aspiration risk, slowing neurological progression, and increasing the lifespan of NPC1 individuals; however, the exact mechanism of action and its impact on the serum proteome remains unknown(12, 13). The FDA has also recently approved N□acetyl□L□leucine (NALL) (Aqneursa®) as a stand-alone treatment for neurological symptoms in both adult and pediatric NPC1 individuals(14). NALL may improve mitochondrial and lysosomal functions by enhancing adenosine triphosphate (ATP) production and normalizing energy metabolism. This may help reduce the pathogenic accumulation of unesterified cholesterol and sphingolipids(14). Identifying informative biomarkers may facilitate diagnosis and prognostic counseling in addition to providing tools to assess the efficacy of therapeutic drugs. However, given the substantial heterogeneity in NPC1 disease onset, severity, and progression, it is essential to account for covariates to ensure that biomarker associations truly reflect disease-specific and treatment-related effects.

In a previous study of cerebrospinal fluid (CSF), we identified and confirmed several protein biomarkers, including PARK7, CALB2/calretinin, CHI3L1/YKL-40, MIF, CCL18, and ENO2, in the CSF of NPC1 individuals compared to non-NPC1 samples(15). This prior study also confirmed proteins previously reported to be increased in NPC1 (NEFL(16, 17), MAPT(18), CHIT1(19), CALB1(20)). Since CSF collection is not a routine procedure in many clinical trials, identifying blood-based biomarkers that correlate with clinical aspects of the disease would be advantageous. In this study, we performed a discovery-based differential proteomic analysis of serum from NPC1 individuals and compared it with healthy pediatric controls. To delineate the effects of NPC1 disease on the serum proteome independent of covariates, we constructed statistical models that accounted for factors potentially influencing protein levels. This approach enabled us to isolate disease-specific proteomic signatures associated with NPC1 pathology. We also assessed the impact of miglustat treatment on the serum proteome to evaluate its potential to reverse disease-associated alterations and to identify candidate biomarkers with clinical relevance for monitoring miglustat response. The study revealed proteins that had not been previously identified as altered in NPC1 individuals and confirmed some previously reported proteins, such as NPY(21), GPNMB(22), and CCL18(15). We confirmed selected differentially abundant proteins using orthogonal assays and explored correlations with clinical parameters to characterize their potential as clinically relevant biomarkers. We identified hundreds of serum proteins with increased abundance in NPC1 disease baseline samples that were then decreased in miglustat-treated samples, thus exhibiting inverse abundance levels between NPC1 disease and miglustat treatment, and could be potential candidates related to the miglustat therapeutic effect.

## Materials and Methods

### Trial participants and biomaterial collection

Individuals with NPC1 disease were enrolled in a natural history/observational trial conducted at the National Institutes of Health Clinical Center (NCT00344331). The *Eunice Kennedy Shriver* National Institute of Child Health and Human Development Institutional Review Board initially approved the study, and the National Institutes of Health Intramural Institutional Review Board has provided the ongoing review. Participants or guardians provided written consent, and assent was obtained when appropriate. Biochemical and molecular testing were used to confirm a clinical diagnosis of NPC1. A total of 68 serum samples from NPC1 individuals and 20 from healthy pediatric controls were obtained and stored at −80°C. The NPC1 cohort included 25 serum samples from individuals without miglustat treatment (disease baseline) and 43 from individuals receiving miglustat treatment. The NPC1 cohort contained eleven paired samples collected from individuals before and after receiving miglustat treatment. Detailed clinical phenotyping of individuals included measuring parameters of disease burden using the 5- and 17-domain NPC Neurological Severity Scores (NSS)(23, 24), as well as disease severity, including Age of Neurological Onset (ANO) and Annual Severity Increment Score (ASIS)(25). The 5-domain NPC NSS (NSS5) (ambulation, fine motor, speech, swallow, and cognition) is clinically important and highly correlated with the complete 17-domain scoring for NPC1. A systematic review of the clinical history was used to determine ANO, and ASIS is calculated by normalizing the 17-domain NPC NSS (NSS17) to the individual’s age at the time of sample collection in years. Earlier ANO and high ASIS are indicative of severe disease. The 5- and 17-domain NPC NSS reflect the current burden of neurological signs/symptoms. The demographics (age and sex) of the NPC1 and control cohorts, as well as the miglustat treatment status of the NPC1 cohort, are provided in Table 1 and Additional Table 1.

**Table 1:** Summary statistics of the NPC1 individuals and the healthy pediatric control cohort.

| Variable | Pediatric Controls | NPC1 | NPC1 No Miglustat | NPC1 Miglustat |
| --- | --- | --- | --- | --- |
| **Screening Cohort** |  |  |  |  |
| Number of Individuals (n) | 20 | 68 | 25 | 43 |
| Mean Age ± SD (years) | 11.7 ± 3.9 | 14.1 ± 13.4 | 16.2 ± 18.5 | 12.9 ± 9.3 |
| p-value | - | 0.64* | - | 0.68<sup>s</sup> |
| Median Age and Range (years) | 11.4 (4.8 - 18) | 9.7 (0.6 - 68.2) | 8.9 (0.6 - 68.2) | 9.8 (0.9 - 35.4) |
| Male/Female (% Male) | 13/7 (65%) | 34/34 (50%) | 12/13 (48%) | 22/21 (51%) |
| **Age of Neurological Onset (years)** |  |  |  |  |
| Number of Individuals (n) | NA | 67<sup>a</sup> | 25 | 42 |
| Mean | | 6.5 ± 8.3 | 7.8 ± 11.8 | 5.7 ± 5.3 |
| p-value |  | - | - | 0.83<sup>s</sup> |
| Median (range) |  | 4 (0.5 - 47) | 4 (0.5 - 47) | 4 (0.8 - 25) |

| NPC 5-domain NSS (points) |  |  |  |  |
| --- | --- | --- | --- | --- |
| Number of Individuals (n) | NA | 58<sup>b</sup> | 21 | 37 |
| Mean |  | 9.3 ± 5.7 | 9 ± 5.7 | 9.5 ± 5.7 |
| p-value |  | - | - | 0.70<sup>s</sup> |
| Median (range) |  | 8.5 (1 - 22) | 8 (1 - 20) | 10 (1 - 22) |
| NPC 17-domain NSS (points) |  |  |  |  |
| Number of Individuals (n) | NA | 65<sup>c</sup> | 23 | 42 |
| Mean |  | 16.2 ± 10.9 | 15.6 ± 9.3 | 16.6 ± 11.8 |
| p-value |  | - | - | 0.83<sup>s</sup> |
| Median (range) |  | 16 (1 - 44) | 17 (1 - 32) | 16 (1 - 44) |
| ASIS (points/year) |  |  |  |  |
| Number of Individuals (n) | NA | 65<sup>c</sup> | 23 | 42 |
| Mean |  | 1.9 ± 2.5 | 1.8 ± 2.2 | 1.9 ± 2.7 |
| p-value |  | - | - | 0.76<sup>s</sup> |
| Median (range) |  | 1.2 (0.07 - 16) | 1.1 (0.15 - 9.4) | 1.2 (0.07 - 16) |

\*Mann-Whitney t-test comparison of the mean age of the control versus the NPC1 group
<sup>s</sup>Mann-Whitney t-test comparison of miglustat-treated and untreated individuals
<sup>a</sup>One individual had no reported neurological symptoms
<sup>b</sup>10 individuals with a 5-domain NPC NSS score = 0
°3 individuals with 17-domain NPC NSS and ASIS = 0

### Proximal extension assay

Serum proteins were analyzed by Proximal Extension Assays (PEA) using the Olink® Explore 3072/384 platform (Olink Proteomics Inc., Boston, MA). NPC1 and control serum samples were randomized on a 96-well plate, sealed and frozen, then shipped overnight on dry ice to Olink®, as per the instructions. The platform measures 2,943 proteins from cardiometabolic, neurology, inflammatory, and oncology panels, providing protein abundance in the form of unit-less, log_2_-transformed Normalized Protein Expression (NPX) values. These NPX values were used for downstream analysis of differentially abundant proteins.

### PEA statistical and bioinformatic analysis

Bioinformatic analyses were performed in R 4.3.3 with packages including OlinkAnalyze 4.2.0 (Nevola et al., 2025), afex 1.4-1 (Singmann et al., 2024), and emmeans 1.11.1 (Lenth, 2025). The code developed for this work is available at https://github.com/NICHD-BSPC/AnalyzeOlink.

#### Quality check

Quantifiable data were obtained for 2,888/2,943 proteins (98%), including NPX values and QC metrics. All observations with Olink® internal “low quality” QC warnings were removed. Additionally, to identify outliers, for each sample, the NPX interquartile range (IQR) was plotted against its median (i.e., a measure of variance versus a measure of magnitude), and Principal Component Analysis (PCA) was performed. Only one sample was excluded, sample NPC91b was a clear outlier in every panel (IQR–median plots) and separated in PCA (Additional Figure 1). In total, 4.1% of the data was removed from either low-quality QC warnings or the single outlier sample.

### Differential abundance analysis

Differentially abundant proteins were identified in two main comparisons described in more detail below: 1) the NPC1 baseline disease effect and 2) the miglustat treatment effect. Given the heterogeneity of NPC1, statistical models were constructed to assess disease-related effects on the serum proteome while adjusting for covariates based on the leave-one-out sensitivity analysis and correlation screening (Additional Methods). The models described below were built independently for each protein. Continuous covariates were mean-centered. All symbols are defined in-line where first mentioned. Subscripts *i* and *j* index the group and subject, respectively. For differential abundance analysis, the models used coefficients and samples as follows:

#### 1) NPC1 baseline disease effect

This model compares untreated NPC1 individuals with healthy controls. The data set excludes all post-miglustat samples *n_NPC_*_1_ = 24, □*n_control_* = 20

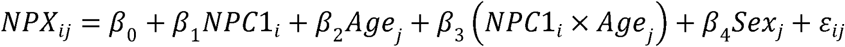

*β*_0_ is the overall mean NPX for the reference sex at the age intercept; *β*_1_*NPC*1*_i_* is the adjusted difference between the NPC1 and control groups, and is the primary coefficient of interest; *β*_2_ *Age_j_* is the change in NPX per year of age; *β*_3_ (*NPC*1*_i_* x *Age_j_*) captures whether the NPC1-control difference varies with age; *β*_4_*Sex_j_* is the mean NPX shift between sexes; and *ε_ij_* is the residual error for subject j.

#### 2) Miglustat treatment effect

This model tests the average difference in NPX between NPC1 individuals who received miglustat and those who did not, using one sample per individual. *n_treated_* = 32, □*n_untreated_* = 24

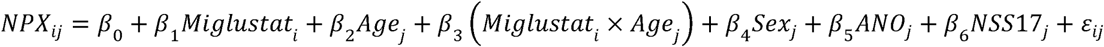

*β*_1_*Miglustat_i_* estimates the adjusted mean NPX difference between treated and untreated NPC1 individuals and is the primary coefficient of interest; *β*_3_ (*Miglustat_i_* x *Age_j_*) captures the age-dependent modification of the treatment effect; *β*_5_ *ANO_j_* is the change in NPX per year of age at neurological symptom onset; and *β*_6_ *NSS*17*_j_* is the change in NPX per point of the 17-domain Neurological Severity Score.

Denominator degrees of freedom were estimated using Satterthwaite’s method (the default for the afex package). Raw p-values for fixed effects were Benjamini–Hochberg–adjusted across proteins (FDR < 0.10). Post-hoc comparisons were obtained with the emmeans package. Since the raw log_2_FC still includes the effects of covariates, we report the “adjusted log_2_FC (Estimate)” which is calculated by emmeans for the main effect of interest, thereby controlling for the effects of covariates.

To identify biomarkers exhibiting inverse expression patterns between the NPC1 baseline disease effect and the miglustat treatment effect (i.e., disease-associated proteins that reverted towards normal after miglustat), we intersected the sets of differentially abundant proteins obtained from the two comparisons. Protein fold changes (adjusted log_2_FC) from both comparisons were plotted against each other to visualize reciprocal expression trends.

### Orthogonal confirmation assays

Human-specific enzyme-linked immunosorbent assays (ELISA) were used to confirm increased expression of TREM2, AgRP, NPY, HSD17B14, GPNMB, Cathepsin L, CCL18, DSCAM, and CEND1. Serum samples were thawed on ice and diluted in sample diluent and assayed in duplicate according to the manufacturer’s protocol as follows: TREM2 (Abcam, ab224881, Waltham MA, USA), AgRP (Human AgRP ELISA kit, ELH-AgRP-1, RayBiotech, Peachtree Corners, GA, USA), NPY (Human Neuropeptide Y/NPY DuoSet ELISA, DY8517-05, R&D Systems, Minneapolis, MN, USA), HSD17B14 (Human HSD17B14 PharmGenie ELISA kit, SBRS0680, Dublin, Ireland), GPNMB (Human Osteoactivin (GPNMB) ELISA kit, EHGPNMB, Invitrogen, Waltham, MA, USA), Cathepsin L (DuoSet ELISA, DY952, R&D Systems), CCL18 (Abcam, ab211649), DSCAM (Human DSCAM ELISA Kit, ELH-DSCAM-1, RayBiotech), and CEND1 (MyBioSource labs, MBS7207588, San Diego, CA, USA). Individual values that were below the limit of detection were set at the lowest limit of detection.

Serum BDNF levels were measured using Simoa™ BDNF discovery kit (Item 102039) on the Quanterix SR-X platform in a 96-well plate format (Quanterix, Billerica, MA, USA). Briefly, the serum was diluted according to the manufacturer’s protocol with sample diluent and assayed in duplicate. The plates were processed using the 2-step digital immunoassay as outlined in the manufacturer’s protocol.

We mirrored the PEA baseline model by replacing NPX with the log_10_ concentration from the ELISA assays, applying the same sample exclusions (no miglustat-treated samples and removal of the outlier sample NPC91b), and using the same covariate structure (age, sex, and the age×NPC1 interaction). Raw p-values across the ELISA targets were Benjamini–Hochberg adjusted with an FDR < 0.1. Graphs were generated using GraphPad Prism 10.3.0.

### Correlation with clinical parameters

We calculated Spearman correlations between PEA NPX values, age of individual at the time of sample collection (Age), ANO, NSS5, NSS17, and ASIS for every protein. Age NPX correlations were calculated using the entire dataset, which included all NPC1 and control samples (*n_NPC_*_1_ = 67 and □*n_control_* = 20). Clinical variable NPX correlations used the NPC1 samples only, since these clinical variables are only recorded for NPC1 individuals (n_NPC1_ = 67). A substantial number of proteins had significant correlations between NPX and disease severity metrics ANO, NSS5, and NSS17. Therefore, we further investigated whether the changes in the serum proteome are influenced by the parameters of disease severity (ANO) and disease burden (NSS17). As NSS17 encompasses and provides greater granularity than NSS5, models were constructed using NSS17 only. The models described below were built independently for each protein. Continuous covariates were mean-centered. All symbols are defined in-line where first mentioned. Subscript *j* indexes the subject.

#### 3) NPC1 age of neurological symptom onset effect

This model tests the association between NPX values and ANO among NPC1 individuals. *n_NPC_*_1_ = 56. Post□treatment samples are excluded to maintain the independence of measurements.

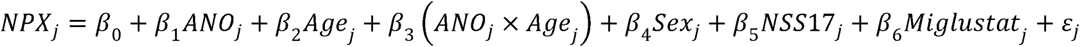

*β*_3_ (*ANO_j_* x *Age_j_*) captures whether the onset age association varies with age at sampling.

#### 4) NPC1 burden effect

This model tests the association between NPX and NSS17 among NPC1 individuals. *n_NPC_*_1_ = 56. Post□treatment samples are excluded to maintain the independence of measurements.

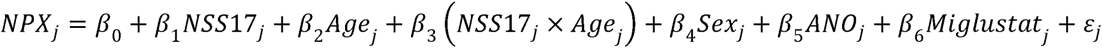

*β*_3_ (*NSS*17*_j_* x *Age_j_*) captures whether the burden association varies with age at sampling.

To evaluate the correlation between clinical parameters and protein concentrations validated by ELISA, Spearman correlations with *rho* values of 0.1-0.3, 0.3-0.5, and >0.5 and p-value < 0.05 were considered weak, moderate, and strong, respectively. Correlation graphs were generated using GraphPad Prism 10.3.0.

### Brain-associated annotations using the Human Protein Atlas

To prioritize serum signals with central nervous system relevance, we annotated each protein using the Human Protein Atlas (HPA; release 24.0, downloaded 2025-08-29). We retrieved the HPA normal-tissue immunohistochemistry (IHC) and tissue-mapping datasets. Brain regions available in the HPA dataset and considered for the analysis were cerebellum, cerebral cortex, hippocampus, and Purkinje neurons. Tissue specificity was quantified as a “tau” specificity score(26) per protein across tissues, which ranges from 0 (broad expression in many tissues) to 1 (tissue-specific expression). Proteins with tau > 0.90 were flagged as brain-specific. IHC categories (“Not detected,” “Low,” “Medium,” “High”) were converted to numeric values 0–3 and summarized into: “Brain Detected” (binary indicator of any detectable expression in a brain region), “Brain Specific” (binary indicator of Tau > 0.90), and “Tau Brain Specificity” (continuous 0–1 Tau score, with 1 indicating expression restricted to brain).

## Results

### Demographic and clinical characteristics of the Niemann-Pick disease, type C1 and healthy pediatric control cohort

Our screening cohort consisted of 68 serum samples from individuals with NPC1 and 20 serum samples from healthy pediatric controls. The average age at the time of sample collection for control samples was 11.7 ± 3.9 years, whereas the average age for NPC1 samples was 14.1 ± 13.4 years (*p* = 0.64). In the NPC1 cohort, the inclusion of older adults’ NPC1 samples (age range: 0.6 – 68.2 years) resulted in a larger standard deviation. The screening cohort consisted of 25 samples from individuals not receiving miglustat treatment and 43 samples collected from individuals receiving miglustat treatment, including some paired, but with no significant difference in the mean age of the two groups (*p* = 0.68). The NPC1 cohort had equal representation of males (50%) and females (50%), with a similar distribution in the miglustat cohort (51% male) and the non-miglustat cohort (48% male). The control cohort consisted of 65% males (*p* = 0.31, Fisher’s exact test). There were no significant differences for any clinical parameters in NPC1 individuals receiving miglustat compared to the untreated group. Demographic details and clinical characteristics of the cohort are provided in Table 1 and Additional Table 1.

### Identification and orthogonal validation of differentially abundant proteins in Niemann-Pick disease, type C1

The relative abundance of serum proteins was determined using the Proximal Extension Assay (PEA, Olink® Explore). Quantifiable data were obtained for 2888/2943 (98%) proteins. When comparing NPC1 individuals at baseline who did not receive miglustat (n = 24) with healthy pediatric control samples (n = 20), we identified a total of 1054 significantly differentially abundant proteins (adj. p-value < 0.1) in samples from individuals with NPC1 disease after controlling for the age and sex of the individuals. Among these 1054 proteins, 186 proteins were increased with adj. log_2_FC ≥ 1.0 and 286 proteins were decreased with adj. log_2_FC ≤ −1.0 in NPC1 samples (Figure 1a, Table 2, and Additional Table 2).

**Figure 1:**
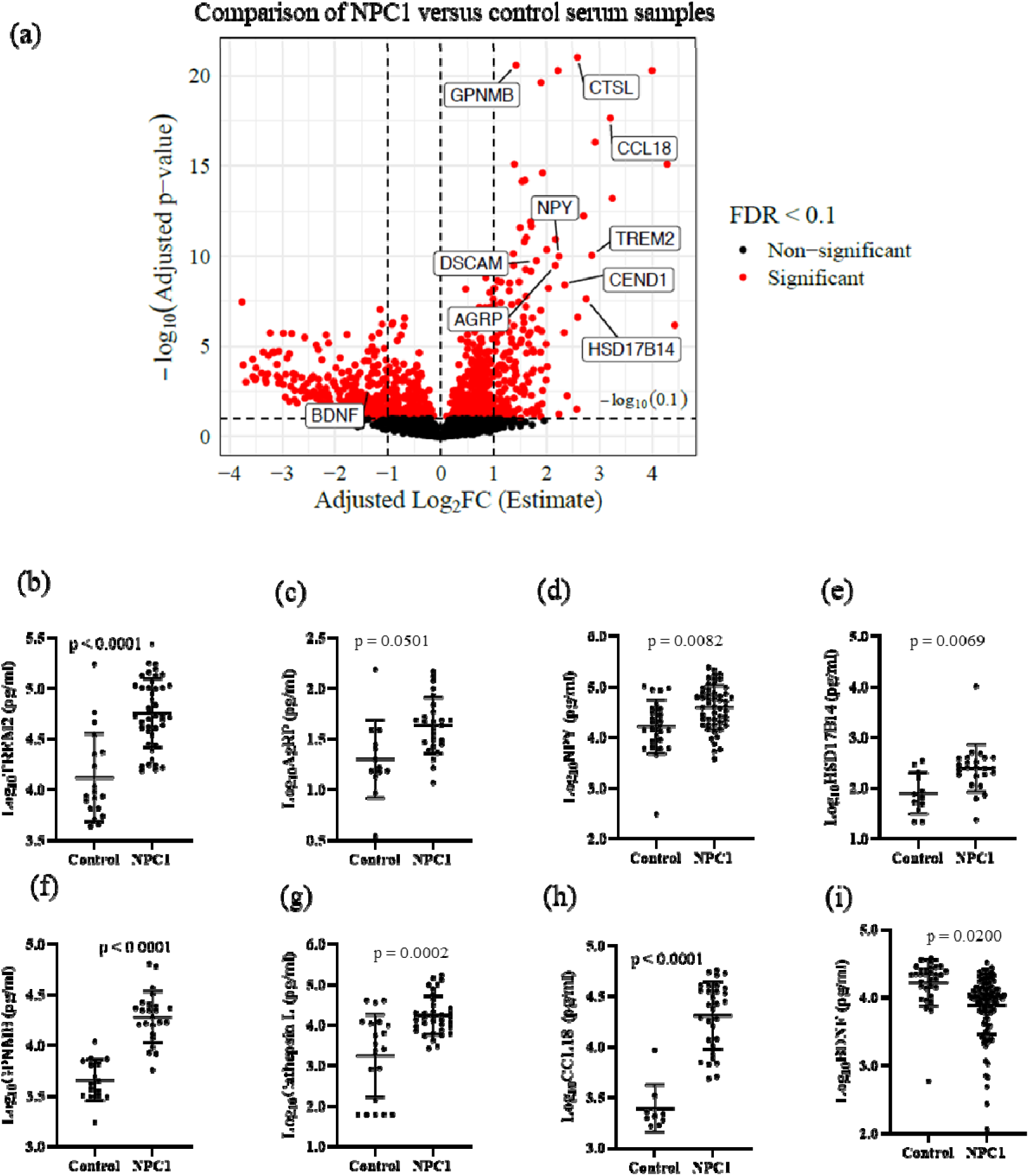
Identification and validation of differentially abundant proteins in NPC1 serum: (a) Volcano plot of differentially abundant proteins comparing NPX values of NPC1 individuals at baseline not receiving miglustat compared to healthy pediatric control, Proteins with adjusted p-value < 0.1 and adjusted log2FC +1 and −1 were considered increased and decreased, respectively. All the proteins with adjusted p-values <0.1 are highlighted in red and proteins selected for validation are labelled. ELISA validation of expression levels of (b) TREM2, (c) AgRP, (d) NPY, (e) HSD17B14, (f) GPNMB, (g) Cathepsin L, (h) CCL18, and (i) BDNF.

**Table 2:**
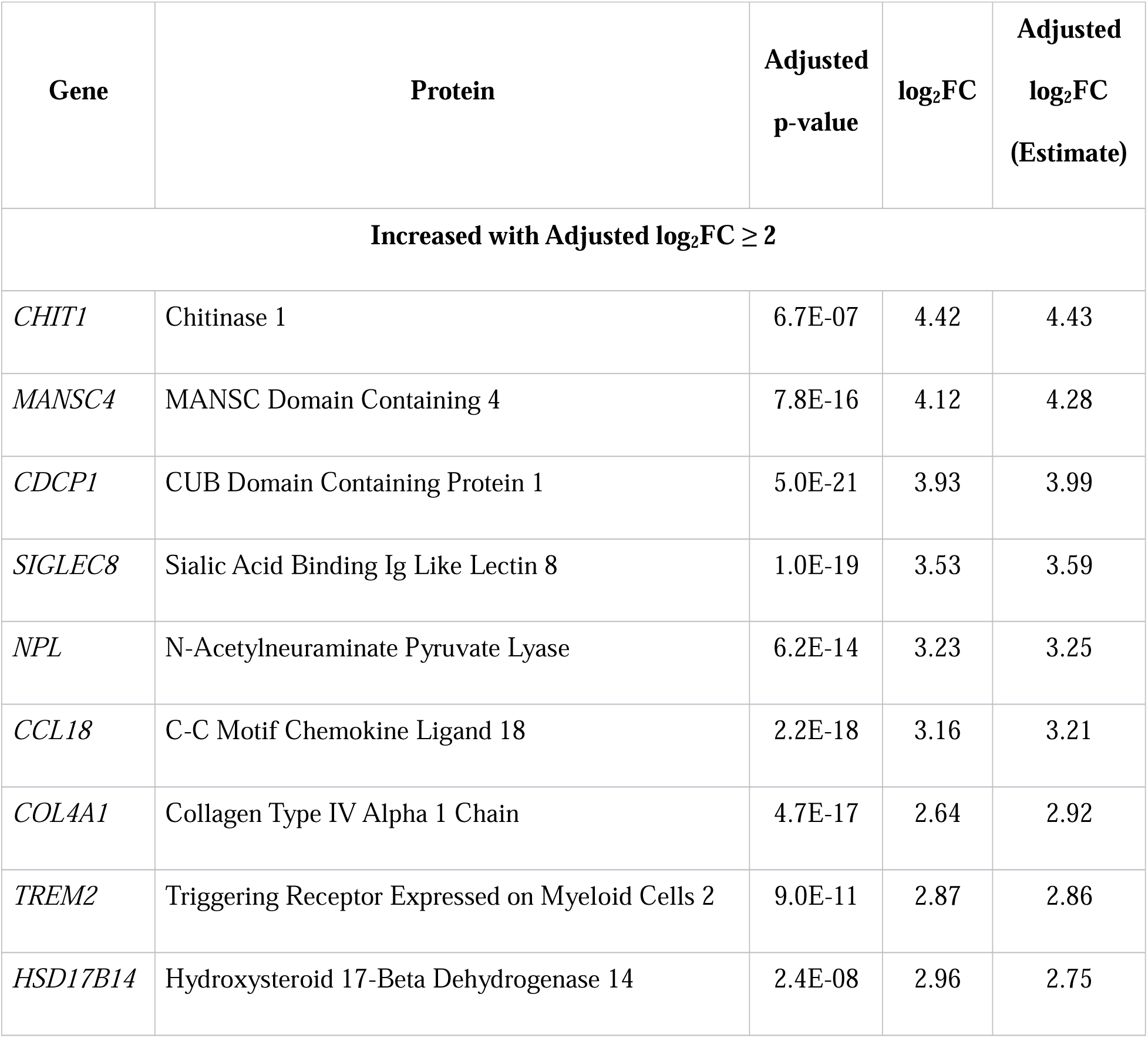
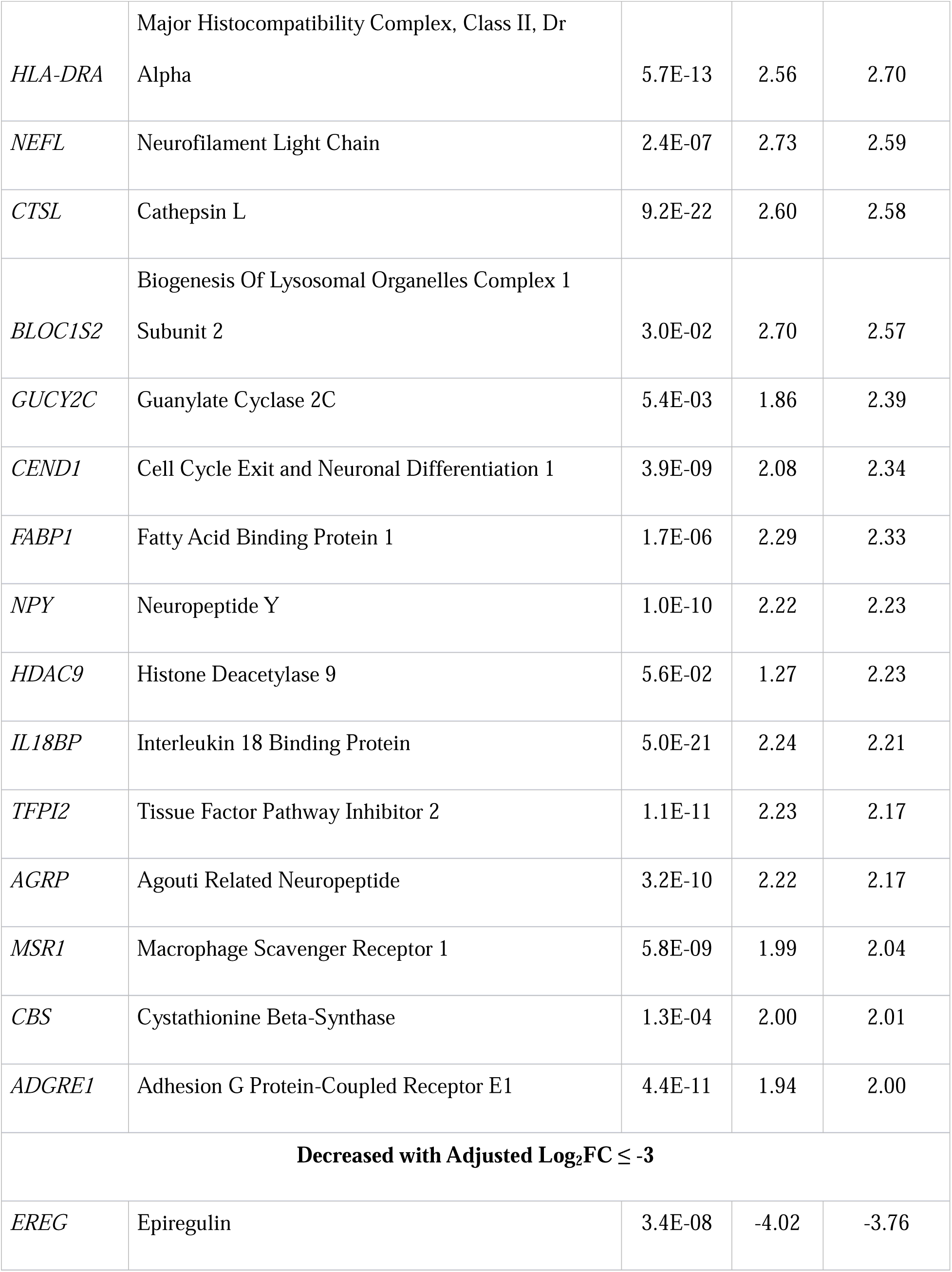
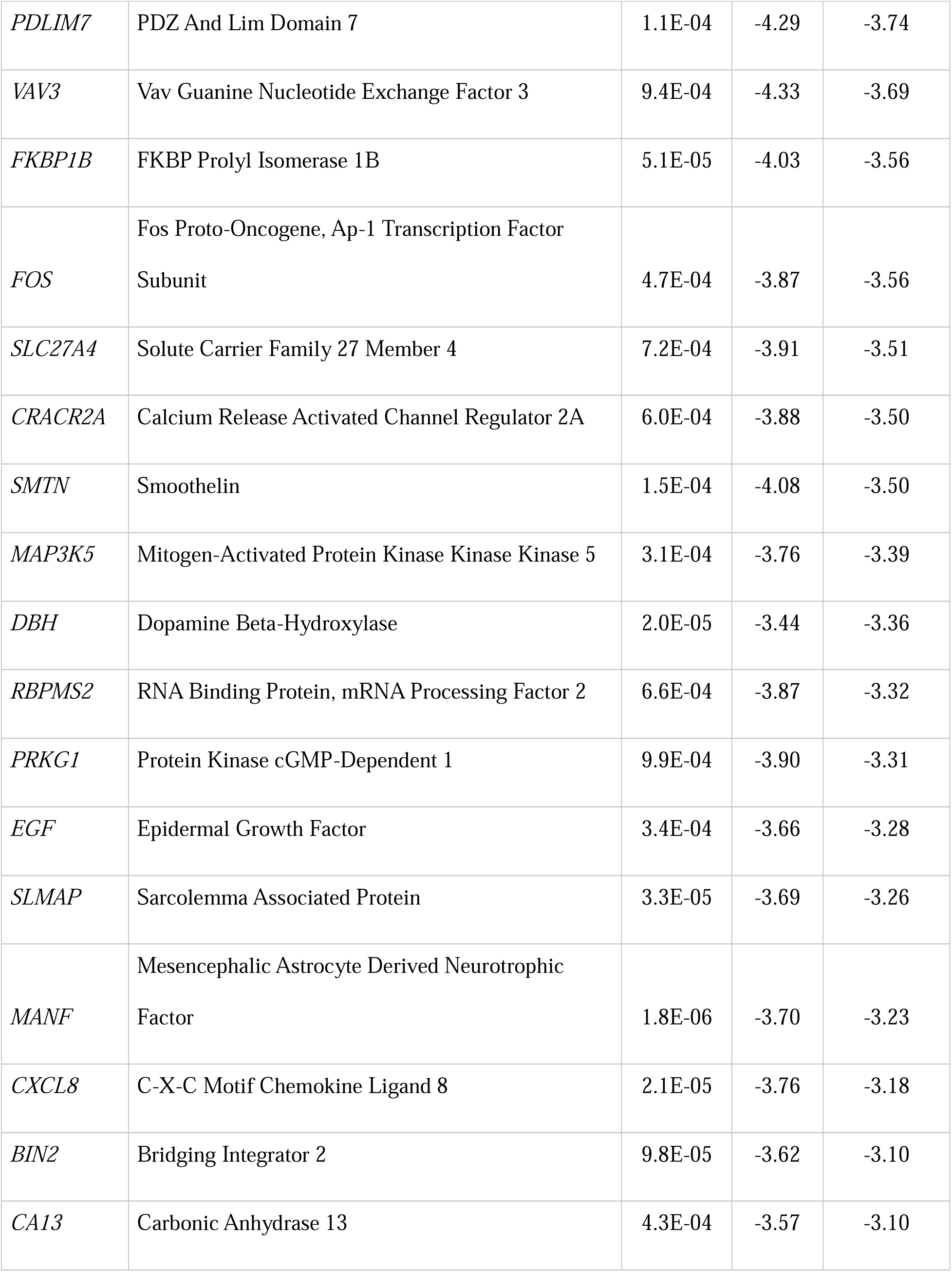
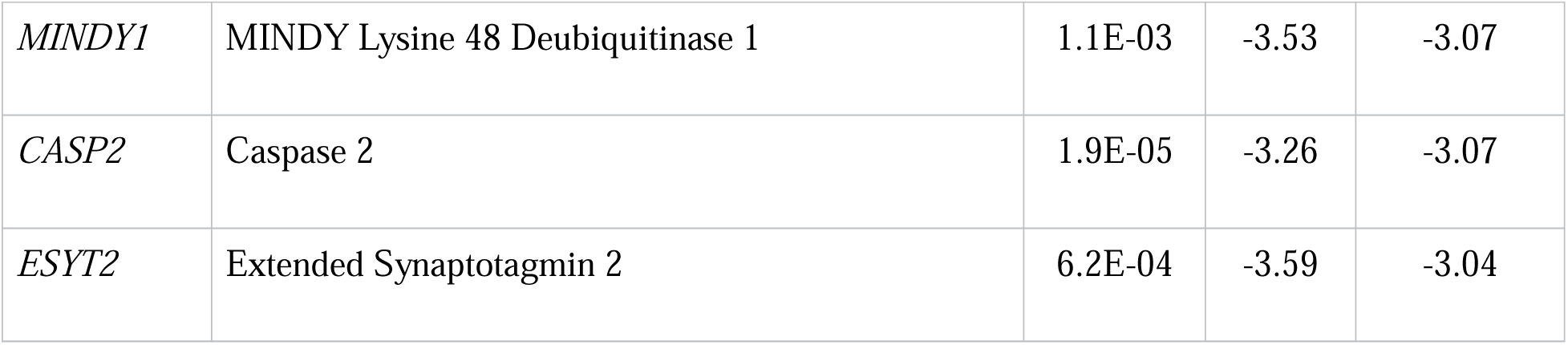
Proteins with increased (log_2_FC ≥ 2) and decreased (Log_2_FC ≤ −3) abundance in NPC1 individual serum samples relative to pediatric controls.

To orthogonally confirm the PEA data, we selected nine proteins that increased and one that decreased in NPC1 individuals for testing by ELISA, as labeled in Figure 1a. We confirmed increased levels of TREM2 (Triggering Receptor Expressed on Myeloid Cells 2), AgRP (Agouti-related protein), NPY (Neuropeptide Y), HSD17B14 (Hydroxysteroid 17-β Dehydrogenase 14), GPNMB (Glycoprotein nonmetastatic melanoma protein B), CTSL (Cathepsin L), and CCL18 (C-C Motif Chemokine Ligand 18), and decreased levels of BDNF (Brain-derived neurotrophic factor) in NPC1 serum samples (Figure 1b – 1i, Table 3). However, we could not validate DSCAM levels and CEND1 expression levels in ELISA assays (Additional Figure 2a, 2b).

**Table 3:**
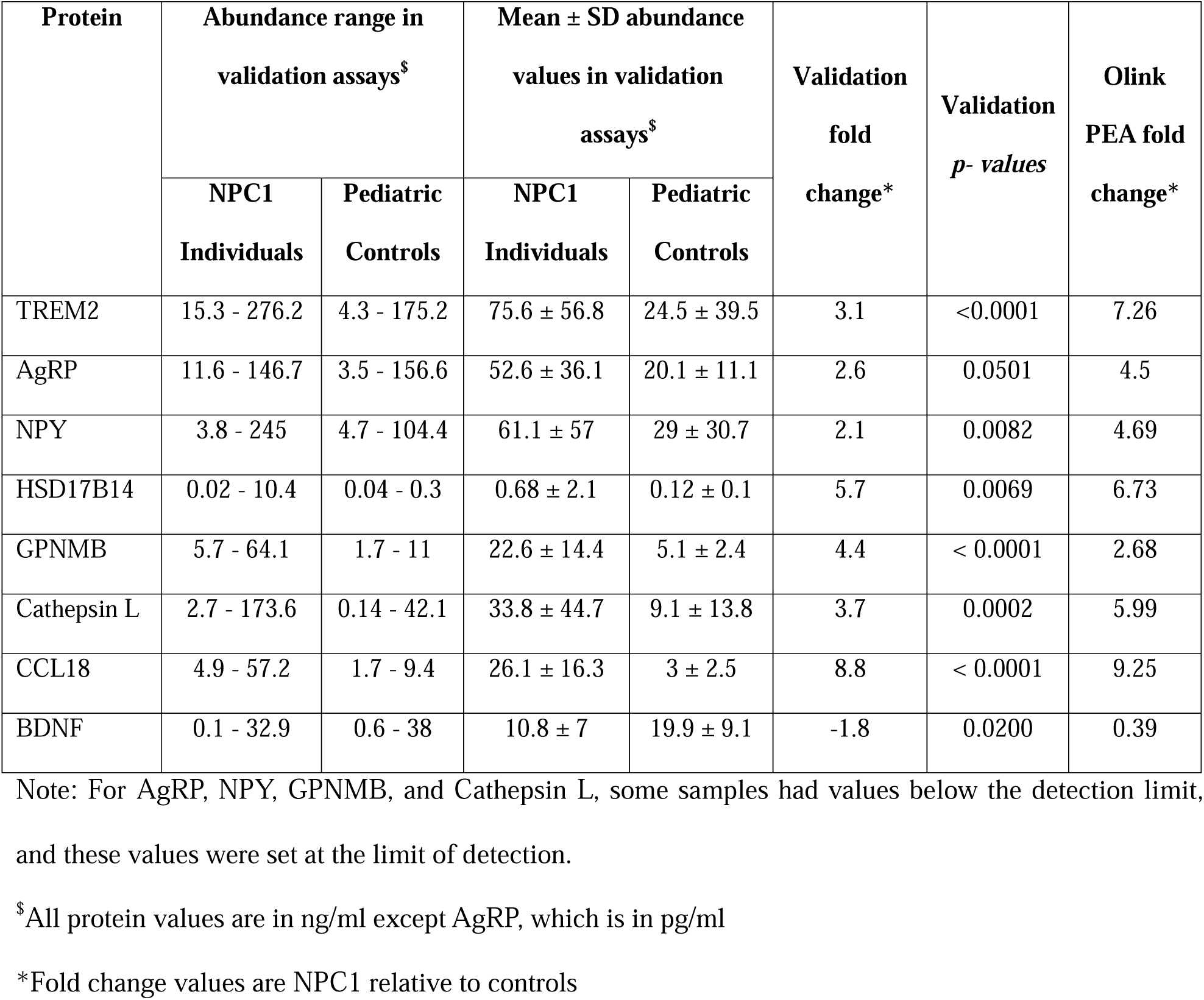
Abundance range and fold change values of serum proteins validated by ELISA assays.

### Effect of miglustat on serum proteins in Niemann-Pick disease type C1

We analyzed the NPX data to identify proteins with altered abundance due to miglustat treatment in NPC1 individuals. We compared unpaired samples from NPC1 individuals receiving miglustat treatment (n = 32) with those of NPC1 individuals not receiving miglustat (n = 24). We identified 197 significantly differentially abundant proteins (adj. p-value < 0.1) due to a miglustat effect, while controlling for age, sex, ANO, and 17-domain NPC NSS of NPC1 individuals. Of these 197 proteins, only four proteins were increased, and the remaining 193 proteins were decreased in the miglustat-treated samples. Interleukin-5 (IL5) and Progestagen Associated Endometrial Protein (PAEP) were increased (adj. log_2_FC ≥ 1.0). Three proteins, namely Meprin A Subunit Alpha (MEP1A), Lacritin (LACRT), and Oxytocin (OXT), were decreased (adj. log2FC ≤ −1.0) in NPC1 individuals receiving miglustat (Figure 2a, Additional Table 3).

**Figure 2:**
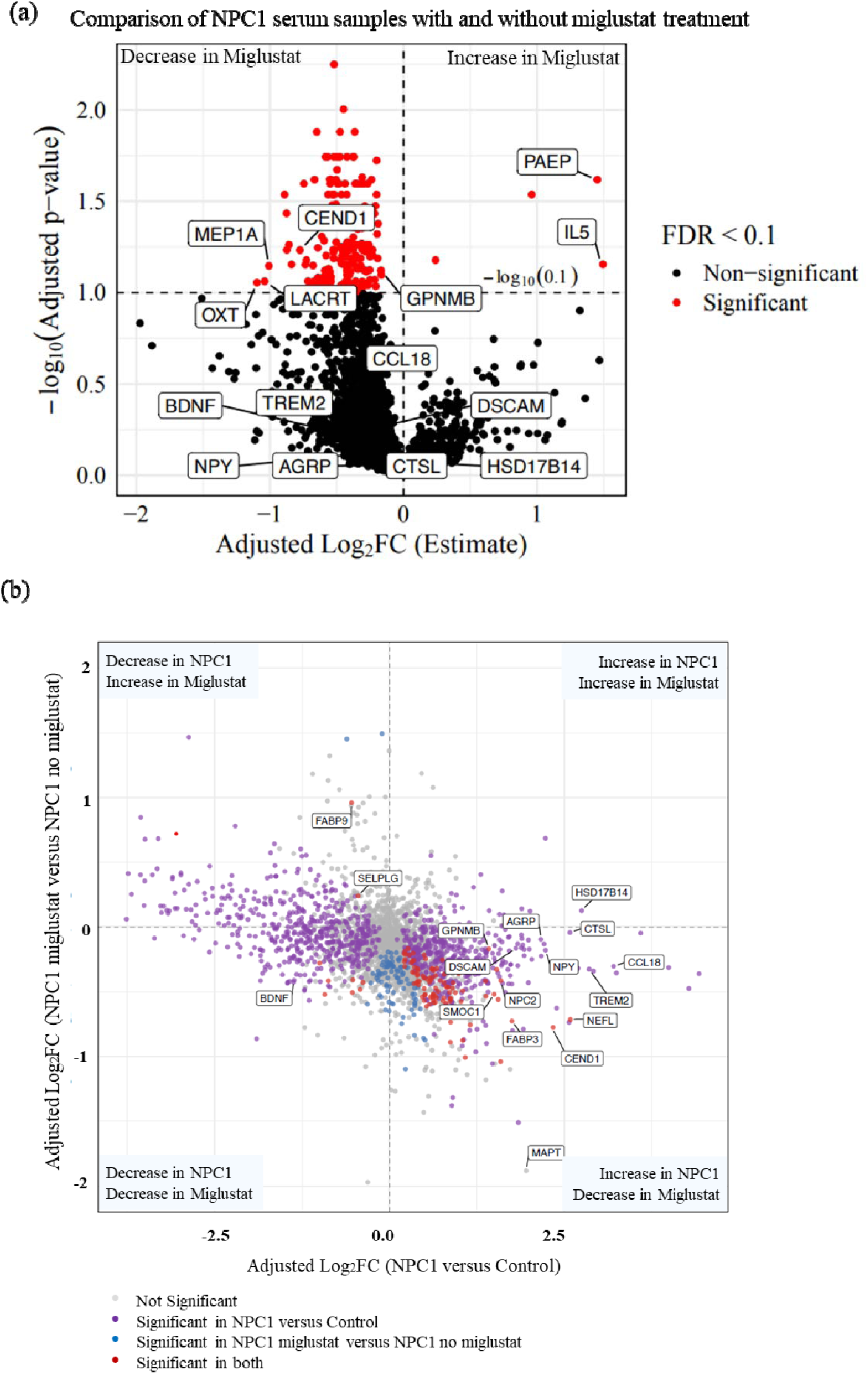
Effect of miglustat on serum proteins in NPC1 individuals: (a) Volcano plot of differentially abundant proteins comparing NPX values of NPC1 individuals receiving miglustat compared to the NPC1 individuals not receiving miglustat. Proteins with adjusted p-value <0.1 and log_2_FC +1 and −1 were considered increased and decreased, respectively. (b) Integration of differentially abundant serum proteins in NPC1 disease, with effect on the X-axis, and miglustat treatment effects on the Y-axis. All the proteins with adjusted p-values < 0.1 are highlighted in red. Proteins discussed in the manuscript are labelled.

We integrated the two distinct lists of differentially abundant serum proteins, first from the NPC1 versus control comparison and second from the miglustat versus non-miglustat comparison, to identify proteins that changed due to NPC1 and shifted back towards control levels with miglustat treatment. Among the 1054 significantly differentially abundant proteins in NPC1, 715 serum proteins showed an inverse expression pattern when compared to miglustat-treated samples. 504 proteins were increased in NPC1 and showed a decrease in expression levels in miglustat-treated samples, and 211 proteins decreased in NPC1 and showed an increase in miglustat-treated samples; however, not all crossed the significance criteria in the miglustat comparison (Additional Table 4). In the NPX data, GPNMB and CEND1 were significantly decreased by 0.89-fold (adj. p-value = 0.0756) and 0.58-fold (adj. p-value = 0.0585), respectively, after miglustat treatment. However, in the ELISA assays, we did not find any significant differences in protein levels due to miglustat treatment (Figure 2a, Additional Figure 3), which could be attributed to the small sample sizes per group in ELISA validations.

We identified 100 proteins that had a significant inverse expression pattern in miglustat-treated samples compared with baseline NPC1 disease. Of these proteins, 98 were increased in NPC1 and were significantly decreased by miglustat treatment. Only two proteins, Fatty Acid Binding Protein 9 (FABP9) and Selectin P Ligand (SELPLG), had decreased levels in NPC1 and were significantly increased by miglustat treatment (Figure 2b, Additional Table 4). Several proteins previously reported to be elevated in NPC1, including MAPT(18), FABP3(27), and NEFL(17), were reduced in miglustat-treated samples. NPC2, the soluble cholesterol-binding partner of NPC1, was significantly increased in serum from individuals with NPC1 disease and showed a marked decrease following miglustat treatment. Notably, secreted modular calcium-binding protein 1 (SMOC1), a recently described early biomarker of Alzheimer disease that is elevated in cerebrospinal fluid years before symptom onset, also exhibited an inverse expression pattern with significant increase in NPC1 disease and significant decrease following miglustat treatment. Although the precise role of SMOC1 in AD remains unclear, it has been shown to colocalize with amyloid-β and influence amyloid aggregation, suggesting potential convergence of neurodegenerative pathways(28).

### Correlation of PEA NPX values with NPC1 clinical parameters

To assess the potential clinical utility of these protein biomarkers, we evaluated Spearman correlations between PEA raw NPX values of all 2888 quantified proteins and phenotypic parameters of NPC1 disease. In total, 1173 (40.6%) proteins significantly correlated with age, of which 163 proteins showed positive correlation, and 1010 showed negative correlation. Thus, underscoring the importance of using age-appropriate control samples and including age as a potential covariate. When the correlation was evaluated between the Age of Neurological Onset (ANO), an assessment of disease severity, we identified 81 (2.8%) proteins that correlated positively and 442 (15.3%) proteins that correlated negatively with the age at onset. Using Annual Severity Increment Score (ASIS) as an additional measure of disease severity, only two proteins, NEFL and CD14, showed significant positive correlations, while ADP-Ribosyl Transferase 3 (ART3) exhibited a significant negative correlation. Interestingly, NEFL and CD14 were significantly negatively correlated with ANO, whereas ART3 was significantly positively correlated with ANO (Additional Table 5). Correlation with the 17-domain NPC-NSS, a measure of current disease burden, revealed 34 (1.2%) proteins that were positively correlated and 380 (13.3%) proteins that were negatively correlated. Analysis of the 5-domain NPC-NSS—a subset of the 17-domain score—identified 36 (1.3%) positive and 362 (12.6%) negative correlations. As expected, the two scales shared overlapping proteins, 66.7% proteins positively correlated, and 82.8% proteins negatively correlated were shared between NSS17 and NSS5 (Additional Figure 4 and Additional Table 6). This supports the use of the 5-domain NPC-NSS as a functional surrogate for the full 17-domain assessment(24, 29).

While the Spearman test provides a robust, non-parametric assessment of associations, it does not fully capture the influence of potential covariates in heterogeneous diseases like NPC1, especially in large sample sizes. To complement these findings, we therefore reanalyzed the data using statistical models that incorporate covariates of age, sex, and miglustat status. Since disease onset age can shape the long-term disease trajectory, we next examined whether age at neurological symptom onset (ANO) was associated with serum proteomic variation while adjusting for age, sex, and miglustat status in the NPC1 individuals. However, no proteins reached the 10% FDR threshold for ANO (Figure 3a, Additional Table 7). Next, we hypothesized that individuals with a higher disease burden would exhibit distinct proteomic changes compared to those with a lower burden. To test this, we incorporated NSS17 as a covariate in the model. Among 56 NPC1 samples, 34 proteins were significantly associated with NSS17 (adj. p-value < 0.1). Of these, 19 proteins increased in abundance, and 15 proteins decreased with increasing NSS17 (Figure 3b, Additional Table 8).

**Figure 3:**
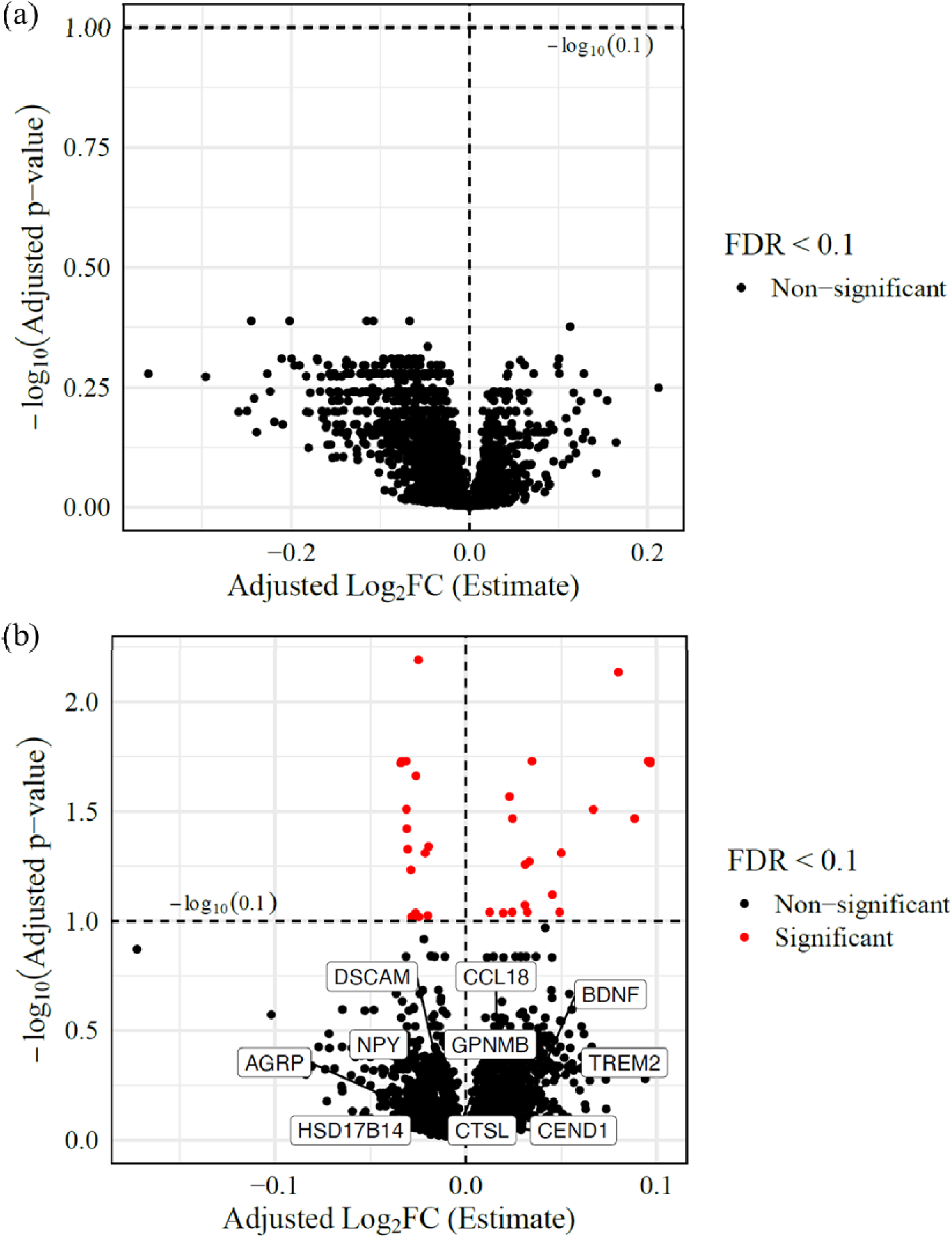
Correlation with clinical parameters: Volcano plot of differentially abundant proteins correlated with (a) ANO and (b) 17-domain NPC NSS. All the proteins with adjusted p-values < 0.1 are highlighted in red. The plot highlights the effect of NSS17 score on protein abundance levels. For instance, with a one-point increase in NSS17 score, IL6 levels will have a significant increase of 0.096 (adjusted Log2FC).

### Correlation of ELISA-validated proteins with NPC1 clinical parameters

Differential protein abundance of TREM2, AgRP, CCL18, Cathepsin L, GPNMB, NPY, HSD17B14, and BDNF was confirmed by analyte-specific ELISA (Figure 1). TREM2, a transmembrane glycoprotein expressed in microglia, is associated with Alzheimer disease and showed a strong negative correlation with the age of neurological onset (rho = −0.475, *p* = 0.0015) (Figure 4a). The correlation value did not change substantially when individuals with an age of neurological onset greater than 20 years were excluded from the analysis (rho = −0.514, *p =* 0.0007) (Additional Figure 5a). TREM2 expression values did not correlate with ASIS, 17-domain NPC NSS, or 5-domain NPC NSS (Additional Figure 5b-5d).

**Figure 4:**
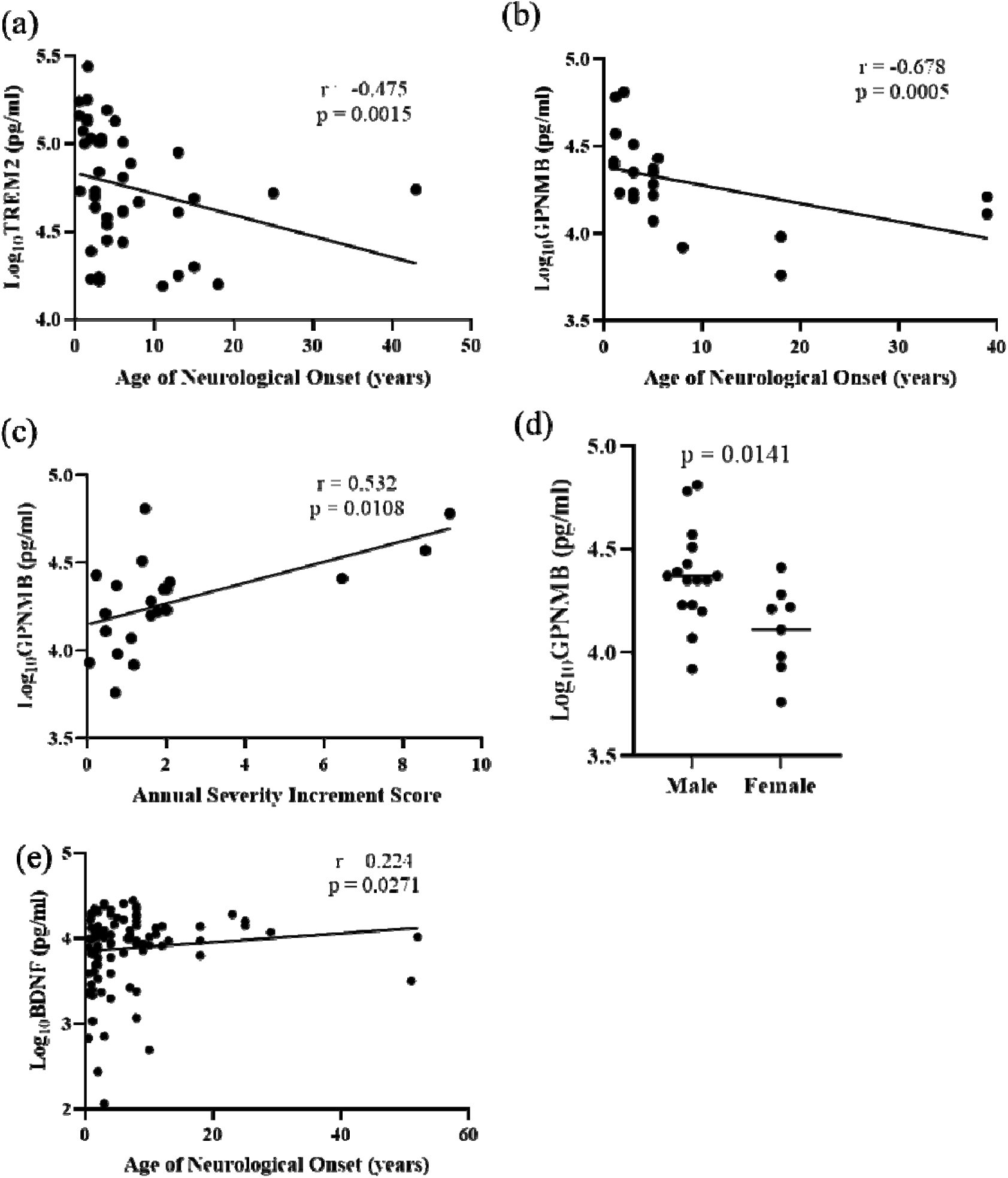
Spearman Correlation of clinical parameters with ELISA (a) TREM with Age of neurological onset, (b) GPNMB with age of neurological onset, (c) GPNMB with Annual Severity Increment score, (d) GPNMB serum levels in male and female NPC1 individuals, and (e) BDNF correlation with Age of neurological onset. Spearman correlations with rho-values of 0.1-0.3, 0.3-0.5, and >0.5 and p-value < 0.05 were considered weak, moderate, and strong, respectively. An unpaired two-tailed t-test was used to evaluate the differences in GPNMB serum levels between males and females.

Levels of GPNMB, a multifunctional protein important for maintaining lysosomal integrity, showed a strong correlation with disease severity parameters. GPNMB was negatively correlated with the age of neurological onset (rho = −0.678, *p =* 0.0005) and positively correlated with the ASIS (rho = 0.532, *p =* 0.0108) (Figure 4b, 4c). GPNMB did not correlate with either the 5- or 17-domain NSS, which are measures of disease burden (Additional Figure 6a and 6b). GPNMB is also reported to be regulated by sex hormones in animal model studies(30). Among the seven proteins tested, only GPNMB showed sex differences present at ∼1.8-fold higher levels (*p =* 0.0141) in males (23.42 ± 15.54 ng/ml) compared to females (12.91 ± 6.5 ng/ml) (Figure 4d, Additional Figure 7).

BDNF is an extensively studied member of the neurotrophin family of growth factors which promotes the growth and differentiation of new neurons and supports the survival of existing neurons(31). Levels of BDNF showed a weak positive correlation with age of neurological onset (rho = 0.224, *p =* 0.0271) (Figure 4e). BDNF levels did not correlate with ASIS, 17-domain NPC NSS, or 5-domain NPC NSS (Additional Figure 8). Among the ELISA-validated proteins, HSD17B14, CCL18, AgRP, NPY, and Cathepsin L levels did not correlate with ANO, ASIS, or NPC NSS (Additional Figures 9, 10, 11, 12, and 13).

## Annotation with the Human Protein Atlas

Since Niemann-Pick disease type C1 (NPC1) is a neurodegenerative disorder, CNS-derived proteins are more likely to capture disease-relevant neurological processes. In contrast, serum protein levels can be influenced by peripheral pathologies, introducing potential confounders. Thus, prioritizing proteins enriched in the CNS improves biological specificity and strengthens the interpretation of our serum-based findings. To address this, we sought to determine how many of the differentially abundant proteins identified in serum samples could reflect underlying brain pathology. To this end, we leveraged data from the Human Protein Atlas (HPA), which provides immunohistochemistry-based protein expression profiles across several brain regions, including the cerebellum, cerebral cortex, hippocampus, and Purkinje neurons. Purkinje neuron-derived proteins are of particular interest because they are particularly vulnerable to degeneration in NPC1, and their loss contributes to cerebellar ataxia. Among the 1,054 proteins differentially abundant in NPC1 serum samples relative to healthy controls, 584 were detected in brain tissue according to HPA datasets. Of these, 325 were increased, and 259 were decreased in NPC1 serum (Figure 5, Additional Figure 14 and 15, and Additional Table 9). Based on the HPA “tau” score, ranging from 0 (broad expression in many tissues) to 1 (tissue-specific expression), 25 of the differentially abundant proteins in serum were classified as brain-specific (tau score > 0.90). Among brain-specific proteins, the NPC1-increased proteins included DSCAM, CEND1, NEFL, NRCAM, MAPRE3, GFAP, CNTNAP2, SYT1, CSPG5, QDPR, KIAA0319, CHRM1, APLP1, NRXN3, MOG, STX1B, SLITRK1, BCAN, and PTPRZ1. Conversely, brain-specific proteins decreased in NPC1 serum, including SERPINI1, PLXNB3, NRGN, BDNF, WASF3, and DLG4 (Additional Table 10, Additional Figures 14, and 15). Notably, BDNF (tau = 0.906), validated in ELISA assays, was designated as brain-specific. Similarly, CEND1 and DSCAM, although not validated by ELISA, also met the criteria for brain specificity using the HPA dataset. TREM2 and HSD17B14 exhibited medium-level expressions across all four brain regions examined, which, in the context of the HPA, indicates a moderate immunohistochemical signal intensity. NEFL (adjusted log_2_FC = 2.59, tau = 0.975), a previously reported biomarker from CSF, showed relatively high expression in the hippocampus and cerebral cortex compared to the cerebellum and Purkinje neurons (Figure 5a)(15). Among the proteins decreased in NPC1 disease, CACYBP (Calcyclin-Binding Protein) showed high expression in all four examined brain regions. CACYBP contributes to neurodevelopmental processes through regulation of cytoskeletal organization and ubiquitin-mediated protein degradation. Another protein of interest, Mesencephalic Astrocyte-Derived Neurotrophic Factor (MANF), exhibited high expression levels in three of the four brain regions investigated, namely the cerebellum, hippocampus, and Purkinje neurons. MANF is a key neurotrophic factor involved in endoplasmic reticulum stress response and protection against neuronal degeneration. Their high levels in the brain and reduced levels detected in NPC1 serum highlight potential disruptions in neuroprotective and developmental pathways, emphasizing their relevance to NPC1 neuropathology and the need for further functional studies to elucidate their roles in disease progression (Figure 5b). In summary, this analysis highlighted the proteins that are altered in serum but could be better indicators of CNS-derived pathologies. Overall, several NPC1-altered proteins exhibited patterns consistent with CNS involvement across multiple brain regions, including the cerebellum, cerebral cortex, hippocampus, and Purkinje neurons, highlighting their potential as peripheral indicators of neurodegenerative pathology in NPC1.

**Figure 5:**
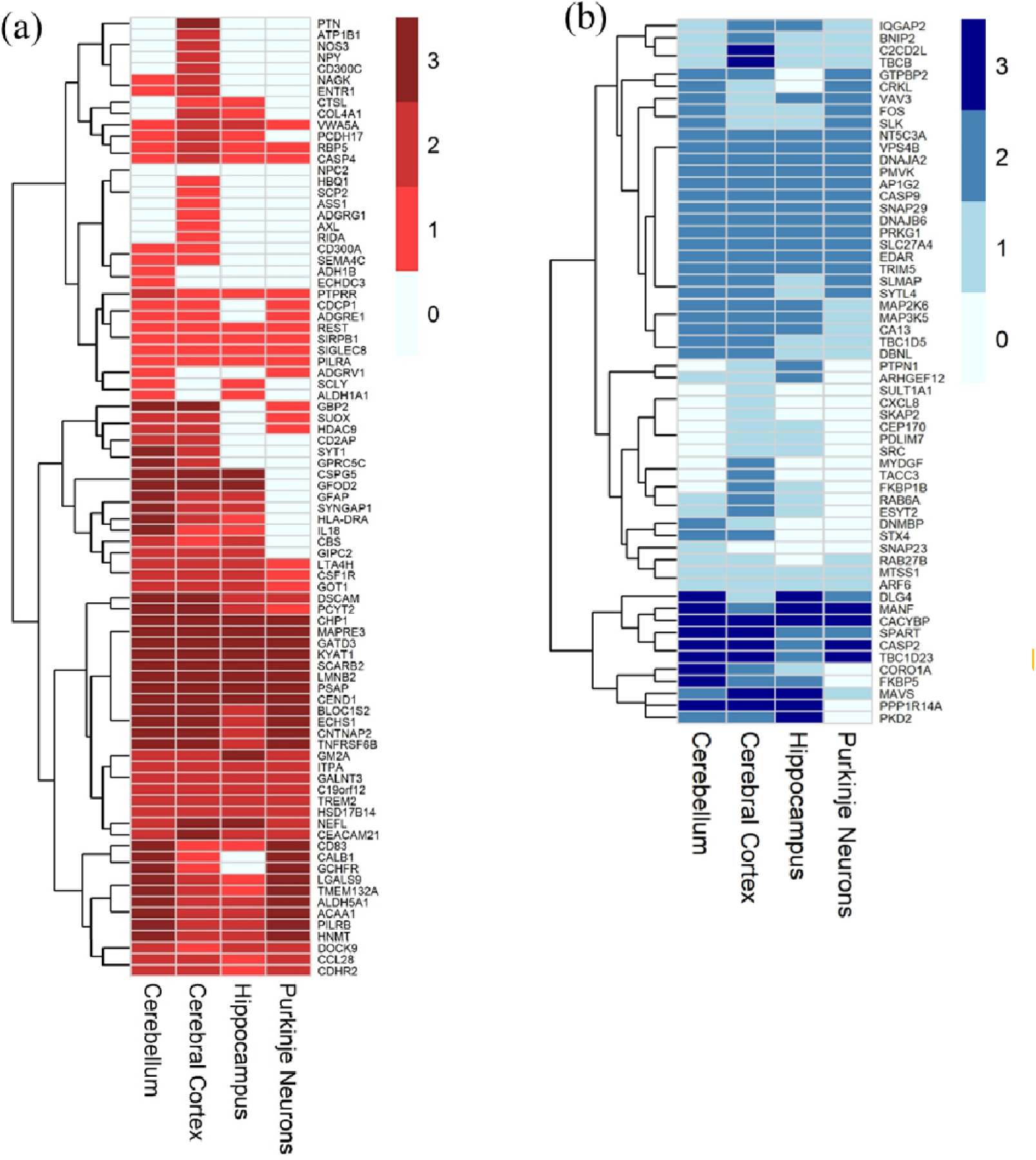
Graphical representation of significantly differentially abundant proteins in NPC1 detected in the Human Protein Atlas (a) Increased in NPC1 with adjusted log2FC ≥1 and (b) Decreased in NPC1 with adjusted log2FC ≤ −2. Scores 0, 1, 2, and 3 represent “not detected”, “low”, “medium”, and “high” expression, respectively, in the HPA dataset.

## Discussion

This study presents a platform-based, high-throughput proteomic approach for identifying potential serum biomarkers in individuals with Niemann-Pick disease type C1 (NPC1). By comparing serum protein profiles from NPC1 individuals and healthy pediatric controls, while adjusting for key covariates, we aimed to identify NPC1 disease–specific alterations. Given the marked heterogeneity of NPC1 in terms of age of onset, severity, and clinical presentation, this analytical framework enabled us to distinguish proteomic changes directly attributable to disease pathology from those influenced by individual variability, thereby providing a clearer molecular signature of NPC1. In this study, we found the increased expression of some previously reported proteins in NPC1 individuals, including NPY, GPNMB, NEFL, MAPT, and CCL18. We extended our knowledge by validating the increased expression of novel proteins such as TREM2, AgRP, HSD17B14, and Cathepsin L, and the decreased expression of BDNF.

We identified elevated levels of Triggering Receptor Expressed on Myeloid Cells 2 (TREM2), a cell surface transmembrane glycoprotein expressed on myeloid cells in multiple organs, including the brain, where it is localized exclusively to microglia. TREM2 forms a signaling complex with the protein tyrosine kinase binding protein (TYROBP) to regulate the immune response by promoting phagocytosis while suppressing cytokine-mediated inflammation. Proteolytic cleavage of its extracellular domain releases soluble TREM2 (sTREM2) into circulation, which has been reported to be elevated in the cerebrospinal fluid of Alzheimer disease (AD) patients and correlated with classical AD biomarkers(32). In NPC1 serum, sTREM2 levels were ∼4.4-fold higher than in controls and negatively correlated with the age of neurological onset, suggesting that higher TREM2 levels may be associated with earlier disease manifestation. Interestingly, while TREM2 mRNA expression increases with age in humans, we observed an age-related decrease in protein levels in NPC1 individuals (rho = –0.475, *p* = 0.0015). Previous studies have demonstrated that TREM2 regulates lipid metabolism and cholesterol turnover in microglia, processes disrupted in NPC1. Elevated sTREM2 may therefore reflect a compensatory response to myelin and lipid abnormalities, or it may contribute to inflammation through decoy receptor activity via sTREM2. Further investigation into the role of TREM2 may provide mechanistic insights into its relationship with the NPC1 disease pathology.

In this study, we found elevated levels of two orexigenic (appetite-inducing) neuropeptides, namely Agouti-related protein (AgRP) and Neuropeptide Y (NPY), in NPC1 individuals. AgRP, primarily expressed in the hypothalamus and adrenal gland, regulates weight homeostasis by controlling feeding behavior by antagonizing melanocortin receptors and modulating intracellular calcium signaling(33). Along with NPY, it increases appetite and decreases metabolism and energy expenditure. Previous studies have associated elevated AgRP with insulin sensitivity and hypothalamic dysfunction, and its increased levels in NPC1 suggest possible metabolic dysregulation(34). Efficient nutrition and monitoring of feeding behavior are important for the care of NPC1 individuals, as they may also develop dysphagia (difficulty in swallowing) that worsens over time and interferes with their feeding habits. Humans and mice affected with NPC1 disease develop metabolic phenotypes, including weight loss, abnormal blood lipid levels, and liver steatosis. NPC1 haploinsufficiency is associated with weight gain, insulin resistance, and obesity in mice and humans, and NPC1 deficiency is associated with an increase in glucocorticoid expression(35). In rats, glucocorticoids increase AgRP and NPY expression in the arcuate nucleus via AMPK signaling, highlighting its importance in maintaining homeostasis.

Neuropeptide Y is one of the most abundant peptides in the nervous system, implicated in neuroprotection and stress regulation. Altered NPY levels have been implicated in multiple neurodegenerative disorders such as Alzheimer disease, Parkinson Disease, and Huntington disease(36). Intranasal administration of NPY in ataxia mouse models has been shown to alleviate the ataxia symptoms without affecting weight gain, food intake, or body fat levels(37). Increased NPY expression in the CSF of NPC1 individuals returns to healthy control levels after miglustat treatment(21), supporting the hypothesis that increased NPY levels might be a self-regulatory mechanism for combating increased neuroinflammation(38). Multiple studies have shown various roles of NPY in modulating neurogenesis, stimulating autophagy, reducing excitotoxicity, and regulating calcium homeostasis(36). In our study, NPY and AgRP did not correlate with any of the clinical parameters; however, their increased expression in NPC1 individuals highlights the need to study these proteins for a better understanding of their role in NPC1, especially in NPC1 heterozygotes, who are more predisposed to obesity(39).

We found increased expression of hydroxysteroid 17-β dehydrogenase 14 (HSD17B14), a key enzyme in the downstream steps of de novo synthesis of neurosteroids and neuroactive steroids, which is known to regulate the relative balance of estrogen and androgen substrates. HSD17B14 regulates sex steroid hormone metabolism and is involved in the final step of sex steroid biosynthesis from cholesterol, using NAD(P)H or NAD(P)+ as cofactors(40). Although involved in the synthesis of sex steroids, no significant differences were observed in males and females in our cohort of NPC1 individuals. It catalyzes the interconversion of estradiol to estrone and testosterone to androstenedione and participates in peroxisomal β-oxidation of fatty acids. Multiple studies have reported the role of HSD17B14 in altered lipid metabolism, steroid synthesis and metabolism, increased reactive oxygen species (ROS), and proinflammatory signaling(41, 42). Studies have shown that levels of steroid hormones such as testosterone, androsterone, progesterone, estrone, and estradiol decreased significantly in NPC1, accompanied by mitochondrial abnormalities, along with increased ROS levels(43). Whether there is any link between the altered steroid levels in NPC1 individuals and increased expression of HSD17B14 remains a subject for future studies.

Glycoprotein nonmetastatic melanoma protein B (GPNMB) is a multifaceted protein that is widely expressed throughout the body, in both transmembrane and secreted form. Studies have reported GPNMB as a potential biomarker for Parkinson disease(44), DM-associated cataracts(45), and Gaucher disease(46). GPNMB mRNA and sGPNMB (soluble GPNMB) levels have been reported to be increased in multiple NPC1 mouse models, as well as in plasma samples from NPC1 individuals(22). In our study, a strong negative correlation was observed between serum GPNMB levels and age of neurological onset. GPNMB levels also showed a positive correlation with annual severity increment scores. These results suggest that increased levels of GPNMB are indicative of severe disease phenotype, and decreasing GPNMB expression might have a positive impact on the attenuation of disease phenotypes. GPNMB expression has been reported to be reduced with intracerebroventricular HP-β-CD treatment along with attenuation of NPC pathophysiology(47). GPNMB was the only protein that showed sex differences, expressed higher in males than females in orthogonally validated assays. In animal model studies, it is regulated by sex hormones, possibly explaining the difference we observed(30). At physiological levels, it is an important protein for maintaining lysosomal integrity, facilitating lysosomal function by aiding phagosome fusion to lysosomes, which leads to efficient macroautophagy in the context of tissue repair. In contrast, its increased expression has been linked to lysosomal stress in Parkinson disease(48).

In this study, we report increased expression of the cytokine C-C Motif Chemokine Ligand 18 (CCL18), a human-specific cytokine secreted by a broad range of monocytes/macrophages and DC antigen-presenting cells of the innate immune system, which primarily affects the adaptive immune systems(49). Increased CCL18 has physiological consequences, and its potential as a biomarker has been reported for numerous diseases, such as glioblastoma(50), Niemann-Pick type B(19), and Gaucher disease(51). A study by Pineda et al.,(52) identified higher CCL18 plasma levels in individuals with either early- or late-infantile onset NPC1, and its higher activity was associated with greater macrophagic activity in early and late-infantile patients versus juveniles, suggesting its potential as an alternative biomarker. Another study suggested that increased plasma CCL18 levels greater than two standard deviations beyond control values should be considered for NPC1 diagnosis(53). Moreover, CCL18 is upregulated due to increased cholesterol accumulation in macrophages(54). In our previous study, we showed increased CCL18 levels in the CSF of NPC1 individuals, a strong negative correlation with age of neurological onset, and a strong positive correlation with ASIS(15). In this study, the CCL18 levels increased in NPC1 serum samples and showed only a moderate correlation with the age of neurological onset. CCL18 belongs to the chemokine family, which, depending on the circumstances, can act as both inflammatory/inducible and constitutive/homeostatic chemokines, thereby complicating its use as a disease monitoring biomarker(55).

Cathepsin L is a cysteine proteinase, with its highest intracellular concentration and activity in the lysosomes. Damage to the lysosomal membranes due to reactive oxygen species has been reported in NPC1 deficiency. It has been shown to mislocalize lysosomal cathepsins due to the leakage of lysosomal contents that culminates in apoptotic cell death(56). This mislocalization is known to affect the activity of cathepsin L, and functional impairments of cathepsin activity have been reported as a feature of NPC1 deficiency(57). Increased cathepsin L expression has been shown in the adipose tissue of NPC2 mice treated with a high-fat diet(58). The protein may play a role in regulating the expression of the oxidized LDL receptor gene, and its loss leads to lysosomal dysfunction(59). The gene has been implicated in many neurodegenerative diseases such as synucleinopathies (Parkinson disease, Dementia with Lewy Body, and Multiple System Atrophy), neuronal ceroid lipofuscinosis, traumatic brain injury, as well as Alzheimer and Huntington disease(60).

We validated the decreased levels of Brain-derived neurotrophic factor (BDNF), an extensively studied member of the neurotrophin family of growth factors. It is involved in encouraging the growth and differentiation of new neurons, supporting the survival of existing neurons, and plays a pivotal role in the learning and memory process(31, 61). BDNF is well studied in neurodegenerative diseases and has been associated with various neuropsychiatric and neurodegenerative disorders like Alzheimer disease, Schizophrenia, Parkinson disease, bipolar disorder, and major depressive disorder(62, 63). Decreased BDNF expression has been reported in serum samples from patients with Alzheimer Disease, diabetic retinopathy(64), plasma samples from heart failure patients(65) and increased BDNF levels have been reported in PTSD patients(66) and epilepsy(67). A study on five NPC1 individuals treated with miglustat reported no change in plasma BDNF levels(68). However, in NPC1 mouse models, a study by Lucarelli et al.(69), have reported dysregulation in the expression and localization of BDNF and its receptor, tropomyosin-related kinase B (TrkB), during early cerebellar postnatal development. These alterations were associated with abnormalities in the differentiation and synaptic connectivity of the granule cells. Another study by Kim et al.(70) reported reduced BDNF expression in the cerebral cortex, brainstem, and cerebellum, during embryonic and early postnatal development of Gaucher disease mouse models, another lysosomal storage disorder. Due to its crucial function in the central nervous system and its altered expression in various neurodegenerative disorders, its utility as a biomarker and therapeutic potential have been evaluated for neurodevelopment(71). Our study, for the first time, reported decreased BDNF serum expression in over 100 serum samples from NPC1 individuals and showed its correlation with the age of neurological onset of the disease.

Both Cell Cycle Exit and Neuronal Differentiation 1 (CEND1) and DS Cell Adhesion Molecule (DSCAM), were increased in the PEA data. CEND1 is a neuron-specific protein located in presynaptic mitochondria, depletion of which can lead to increased mitochondrial fission(72). It plays a critical role during neurogenesis and is necessary for the generation of GABAergic neurons. CEND1 deficiency leads to cognitive impairments in mice, which can be rescued by its over-expression(73). CEND1was highlighted in annotations with HPA datasets showing high expression in all four regions examined and was significantly decreased by miglustat treatment in the NPX data. DSCAM is a cell adhesion protein that plays a pivotal role in neural development and is highly expressed in the developing nervous system(74). It is associated with Down syndrome and has also been shown to be a strong autism risk gene, wherein deficiency of the protein can lead to autism-like behaviors and excessive spine maturation(75). Recently, a study has shown that in the developing cerebellum, DSCAM controls synaptogenesis in Purkinje cells via intercellular association with glutamate-aspartate transporter (GLAST). Although these two proteins are of potentially significant interest with respect to a possible role in NPC1 neuropathology, we were unable to orthogonally validate them with an ELISA assay. This may be due to either different antibodies being used in the two assays or the relatively low number of samples studied by ELISA. Our PEA data suggest that future work investigating these proteins is warranted.

Another key finding of this study is the effect of miglustat treatment on serum protein levels in NPC1 individuals. Miglustat, a glycosphingolipid synthesis inhibitor, is known to alleviate disease symptoms and slow neurodegenerative progression in NPC1. In our dataset, proteins elevated in NPC1 serum, such as TREM2, AgRP, NPY, GPNMB, CCL18, Cathepsin L, HSD17B14, DSCAM, and CEND1, showed reduced abundance following miglustat treatment, approaching healthy control levels, although only a few reached statistical significance. Similarly, the decreased BDNF levels observed in NPC1 individuals increased toward control levels after treatment. Overall, most differentially abundant proteins in NPC1 displayed an inverse expression pattern following miglustat treatment, suggesting that miglustat may partially normalize NPC1-associated molecular alterations, reflecting partial biochemical correction.

## Study limitations

Biomarkers identified from larger sample sizes are generally more reliable; however, due to the rarity of NPC1, our cohort size was limited. Expanding the sample set would enhance biomarker confidence. Serum protein levels may also reflect contributions from peripheral organs rather than the brain, though serum’s accessibility makes it a valuable source. The PEA platform analyzed 2,943 analytes, which limits its coverage compared to whole-proteomics approaches that could reveal additional candidates. As post-translational modifications and proteins below detection limits were not captured, orthogonal validation primarily focused on increased proteins, with only one decreased protein being validated.

## Future Directions

This study identified numerous differentially expressed proteins, though only a few were validated. Many miglustat-altered, brain-expressed proteins warrant orthogonal validation to uncover additional biomarker candidates. Whether these protein changes are causes or consequences of NPC1 remains unknown. Considering the rarity and heterogeneity of the disease, it is important to identify the mechanistic interactions of the validated proteins with NPC1. These proteins can act as genetic modifiers, and interacting proteins might be able to help better understand the heterogeneous nature of the disease severity and phenotypes.

## Conclusions

In this study, we used a proximal extension assay and a statistical model to identify differentially abundant proteins in serum samples from NPC1 individuals compared to healthy pediatric controls, while controlling for covariates. We confirmed the ability of our screen to identify differentially expressed proteins using an orthogonal approach for TREM2, AgRP, NPY, Cathepsin L, CCL18, GPNMB, HSD17B14, and BDNF in the NPC1 individuals. Of these, TREM2, AgRP, Cathepsin L, and HSD17B14 are novel proteins reported in the context of NPC1. NPY, CCL18, and GPNMB have been previously reported to have increased in the CSF of NPC1 individuals, and we showed their increase in serum samples. Our study identified a plethora of differentially abundant proteins that can be selected for future validation and, for the first time, showed the effect of miglustat on the serum proteome. This study is the first approach to identify potential serum protein biomarkers for NPC1 disease that can be used for prognosis or disease monitoring.

## Data Availability

The datasets supporting the conclusions of this article are included within the article and its additional files. Anonymized or coded clinical data is available for IRB-approved research related to NPC1 upon request.

https://github.com/NICHD-BSPC/AnalyzeOlink

## Abbreviations

ASIS: Annual Severity Increment Score
ANO: Age of Neurological Onset
CSF: Cerebrospinal Fluid
ELISA: Enzyme-Linked Immunosorbent Assay
EMA: European Medicines Agency
FC: Fold Change
FDA: Food and Drug Administration
FDR: False Discovery Rate
NPC: Niemann-Pick disease, type C
NPC1: Niemann-Pick disease, type C1
NPC2: Niemann-Pick disease, type C2
NPX: Normalized Protein Expression
NSS: Neurological Severity Score
PEA: Proximal Extension Assay

## Declarations

## Ethics approval and consent to participate

Clinical trial was conducted in accordance with the Declaration of Helsinki. This clinical study was initially approved by the *Eunice Kennedy Shriver* National Institute of Child Health and Human Development Institutional Review Board, and ongoing review has been provided by the National Institutes of Health Intramural Institutional Review Board. Written consent for participation, and when possible and appropriate, assent was obtained.

## Consent for publication

Not applicable.

## Availability of data and materials

The datasets supporting the conclusions of this article are included within the article and its additional files. Anonymized or coded clinical data is available for IRB-approved research related to NPC1 upon request. The code generated for the study is available at https://github.com/NICHD-BSPC/AnalyzeOlink

## Author contribution

Experiment Design: NXC, FDP. Clinical Protocol Support and collection of samples: NYF, DA. Performed experiments: KS, NXC. Bioinformatic Analysis: MM, KC, RD. Data Analysis and Interpretation: KS, NXC, FDP, MM, RD. Supervision NXC, FDP, RD. Manuscript writing and editing: KS. Manuscript editing/review: FDP, NXC. All the authors read and approved the final manuscript.

## Acknowledgements

We would like to acknowledge the individual participants and their guardians for their invaluable contribution to this study. This work would not have been possible without their commitment and willingness to participate in research aimed at addressing this devastating disease.

