## Supplementary Materials and Methods for "Identification of serum protein biomarkers in individuals with Niemann-Pick disease, type C1"

### Supplementary Methods *Model diagnostics*

ANO and NSS17 were not used in NPC1 vs. control since these metrics were not captured for control samples. From our metadata correlations analysis (described below), we know age varies with NPX for 1,236/2,861 proteins at a rate of as much as +/-0.3 NPX per year of age. Our sensitivity analysis (described below) shows that age and NSS17 substantially affect our models (partial η²). ANO had a smaller partial η² than age or NSS17, and a strong enough effect to warrant the term’s inclusion in the miglustat-effect contrasts.

We examined between-group variance homogeneity with per-protein pairwise F-tests comparing our contrast arms. Heteroscedasticity was found in 5.6 % of proteins in the NPC1 vs. control contrast and 4.6 % in the miglustat treatment contrast (FDR < 0.10). The unequal-group variance assumption violation was discovered in a negligible proportion of our tests.

We assessed the normality of residuals (normal error) per protein with Shapiro–Wilk tests (Öztuna, 2006) adjusted by the Benjamini–Hochberg procedure (FDR < 0.10) and confirmed visually with quantile-quantile (QQ) plots. Residual sets with Shapiro–Wilk FDR < 0.10 showed heavy tails and some outliers. Heavy-tailed residuals were detected in 35 % of proteins in the NPC1 vs. control contrast and 43 % for the miglustat contrast.

We sought biomarkers that generalize across the heterogeneous NPC1 population. Classical parametric tests (e.g., ANOVA F-test) are known to become conservative when the error distribution contains biologically driven outliers. The realized Type I error is typically below the nominal α, while power is sacrificed as a tradeoff. This inherent conservatism aligns with our goal of minimizing false-positive protein discoveries in the presence of NPC1 patient-specific idiosyncrasies.

***Sensitivity analyses***

We used Cook’s distance heatmaps and leave-one-out (LOO) diagnostics to quantify the leverage of individual samples and assess each term's influence in our models. For every LOO iteration, we computed partial η² effect sizes with and without a sample or term and visualized the effect of removal with partial η² scatter plots. We inspected samples that appeared frequently or extremely in the heavy tails of the residual distributions. In the NPC1 vs. control contrast, samples showing the most considerable influence contributed to heavy-tail residuals for only a few percent of proteins. We observed similar heavy-tail patterns in the miglustat contrast, but no single sample exerted extreme influence ($\Delta$partial η²) enough to warrant exclusion.

***Covariate balance and metadata correlations***

We checked group comparability with Wilcoxon rank-sum tests for age and χ² tests for sex; no imbalance was detected (all BH-adjusted p ≥ 0.59). We calculated Spearman correlations between NPX, age, ANO, NSS5, NSS17, and ASIS for every protein, controlling the false-discovery rate at 0.10 (Supplementary Figure S1; Supplementary Table S7). Age NPX correlations testing used the full dataset containing all NPC1 and control samples ($n_{NPC1}$ = 67 &  $n_{control}$ = 20). Clinical variable NPX correlations testing used the NPC1 samples only, since these clinical variables are only recorded for NPC1 patients ($n_{NPC1}$ = 67).

Leave-one-out sensitivity analysis and correlation screening showed that age is a major driver: it correlated significantly with 1,173 proteins (FDR < 0.10) and strongly impacted partial η² in our sensitivity analysis. Sex showed lesser partial η² influence, but because sex distributions compared across contrast arms were not perfectly balanced and we have sex linked proteins in our data, we included sex in our models.

Disease severity metrics ANO, NSS5, and NSS17 showed substantial NPX correlations. To avoid over-parameterization in our miglustat effect contrasts, we chose the more granular NSS17 rather than including both NSS17 and the coarser NSS5 to allow due variance to be attributed to severity differences while detecting treatment effects. ASIS, defined as NSS17 divided by age, showed few correlations (1 at FDR < 0.10) and is intrinsically noisy and collinear with NSS17 and age and we therefore omitted ASIS as a covariate in all models.

**Legends Supplementary Tables and Figures**

**Supplementary Tables**

Supplementary Table 1 (ST1): Sample IDs, miglustat status, age, and sex information for the NPC1 individuals and healthy pediatric controls used in the PEA screening. Sample IDs starts with ‘NPC’ and ‘C’ for NPC1 individuals and healthy pediatric control samples, respectively.

Supplementary Table 2 (ST2): Differentially abundant proteins in NPC1 versus control comparisons.

Supplementary Table 3 (ST3): Differentially abundant proteins in NPC1 samples receiving miglustat versus NPC1 samples not receiving miglustat.

Supplementary Table 4 (ST4): Integration of NPC1 disease and miglustat treatment effects on the serum proteome.

Supplementary Table 5 (ST5): Spearman correlation of NPC1 clinical parameters with proximal extension assay NPX values

Supplementary Table 6 (ST6): List of overlapping protein between NSS17 and NSS5; 28 proteins positively correlated with both NSS17 and NSS5 and 336 proteins negatively correlated with both NSS17 and NSS5

Supplementary Table 7 (ST7): Correlation of NPX values with the age of neurological onset with statistical models

Supplementary Table 8 (ST8): Correlation of NPX values with the 17-domain NPC NSS with statistical models

Supplementary Table 9 (ST9): Annotations of differentially abundant proteins in NPC1 with the human protein atlas.

Supplementary Table 10 (ST10): List of differentially altered proteins in NPC1 with brain specific expression (Tau >0.90)

**Supplementary Figures**

Supplementary Figure 1: PCA plot based on NPX values to examine the clustering of samples and identify any potential outliers. Only one NPC1 individual, NPC91b a 29.7-year-old male was an outlier and therefore removed from the downstream analysis.

Supplementary Figure 2 ELISA measurements from NPC1 and control samples (a) GPNMB levels in NPC1 plasma samples (b) DSCAM levels in serum samples, and (c) CEND1 levels in serum samples

Supplementary Figure 3: Expression levels of proteins based on miglustat treatment ELISA assays: there were no differences in the expression level of (a) TREM2, (b) HSD17B14, (c) GPNMB, (d) NPY, (e) Cathepsin L, (f) CCL18, (g) AgRP, (h) BDNF, (i) DSCAM, and (j) CEND1.

Supplementary Figure 4: Overlapping protein correlations between NSS17 and NSS5 (a) Overlap between positively correlated proteins, and (b) Overlap between negatively correlated proteins

Supplementary Figure 5: Spearman correlation of NPC1 serum TREM 2 levels with NPC1 clinical phenotypes (a) age of neurological onset, >20 years excluded from the analysis. (b) annual severity increment score, (c) 17-domain NPC neurological severity score, and (d) 5 domain NPC neurological severity score.

Supplementary Figure 6: Spearman correlation of NPC1 serum GPNMB levels with NPC1 clinical phenotypes (a) 17-domain NPC neurological severity score, and (b) 5 domain NPC neurological severity score.

Supplementary Figure 7: Expression levels of proteins based on gender: there were no gender-based differences in the expression level of (a) TREM2, (b) HSD17B14, (c) NPY, (d) Cathepsin L, (e) CCL18, (f) AgRP, and (g) BDNF.

Supplementary Figure 8: Spearman correlation of NPC1 serum BDNF levels with NPC1 clinical phenotypes (a) annual severity increment score, (b) 17-domain NPC neurological severity score, and (c) 5 domain NPC neurological severity score.

Supplementary Figure 9: Spearman correlation of NPC1 serum HSD17B14 levels with NPC1 clinical phenotypes (a) age of neurological onset (b) age of neurological onset, >20 years excluded from the analysis. (c) annual severity increment score, (d) 17-domain NPC neurological severity score, and (e) 5 domain NPC neurological severity score.

Supplementary Figure 10: Spearman correlation of NPC1 serum CCL18 levels with NPC1 clinical phenotypes Serum CCL18 levels did not correlate with (a) age of neurological onset, (b) Individuals with age of neurological onset >20 years excluded from the analysis. (c) Annual severity increment score, (d) 17-domain NPC neurological severity score, and (e) 5-domain NPC neurological severity score.

Supplementary Figure 11: Spearman correlation of NPC1 serum AgRP levels with NPC1 clinical phenotypes Serum AgRP levels did not correlate with (a) age of neurological onset, (b) Individuals with age of neurological onset >20 years excluded from the analysis. (c) Annual severity increment score, (d) 17-domain NPC neurological severity score, and (e) 5 domain NPC neurological severity score.

Supplementary Figure 12: Spearman correlation of NPC1 serum NPY levels with NPC1 clinical phenotypes Serum NPY levels did not correlate with (a) age of neurological onset, (b) Individuals with age of neurological onset >20 years excluded from the analysis. (c) Annual severity increment score, (d) 17-domain NPC neurological severity score, and (e) 5 domain NPC neurological severity score.

Supplementary Figure 13: Spearman correlation of NPC1 serum Cathepsin L levels with NPC1 clinical phenotypes Serum Cathepsin L levels did not correlate with (a) age of neurological onset, (b) Individuals with age of neurological onset >20 years excluded from the analysis. (c) Annual severity increment score, (d) 17-domain NPC neurological severity score, and (e) 5 domain NPC neurological severity score.

Supplementary Figure 14: Graphical representation of the annotation of significantly increased proteins in NPC1 detected in the Human Protein atlas (a) Increased with adjusted log2FC > = 0.5 and <1 and (b) Increased with adjusted log2FC <0.5

Supplementary Figure 15: Graphical representation of the annotation of significantly decreased proteins in NPC1 detected in the Human Protein atlas (a) decreased with adjusted log2FC <= 1.5 and > -2, (b) decreased with adjusted log2FC <=-1 and >-1.5, (c)decreased with adjusted log2FC <=-0.5 and >- 1, and (d) decreased with adjusted log2FC >- 0.5
