## Supplementary Figures for "Identification of serum protein biomarkers in individuals with Niemann-Pick disease, type C1"

### Slide 1
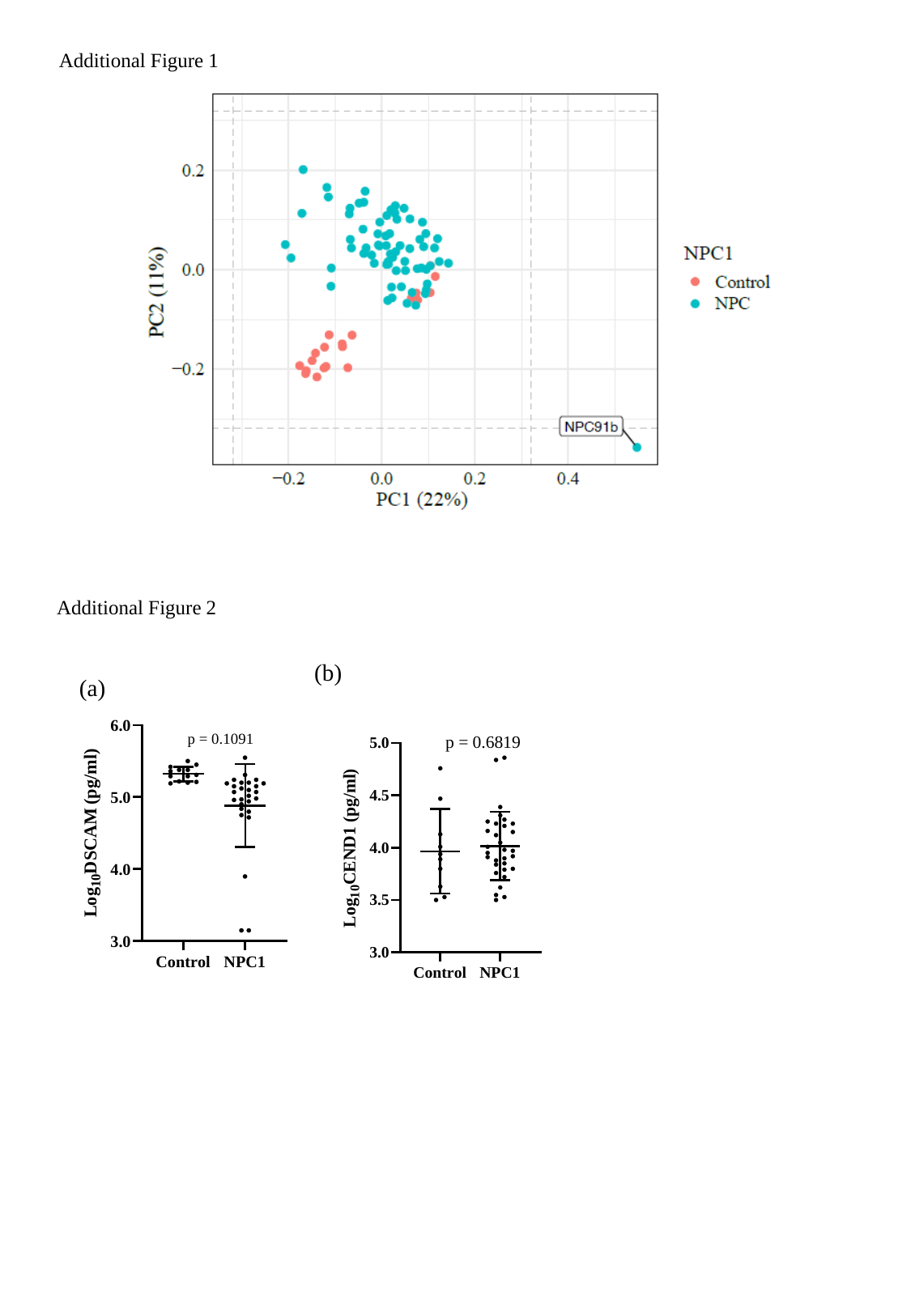

Additional Figure 1
Additional Figure 2
(b)
(a)
p = 0.1091

### Slide 2
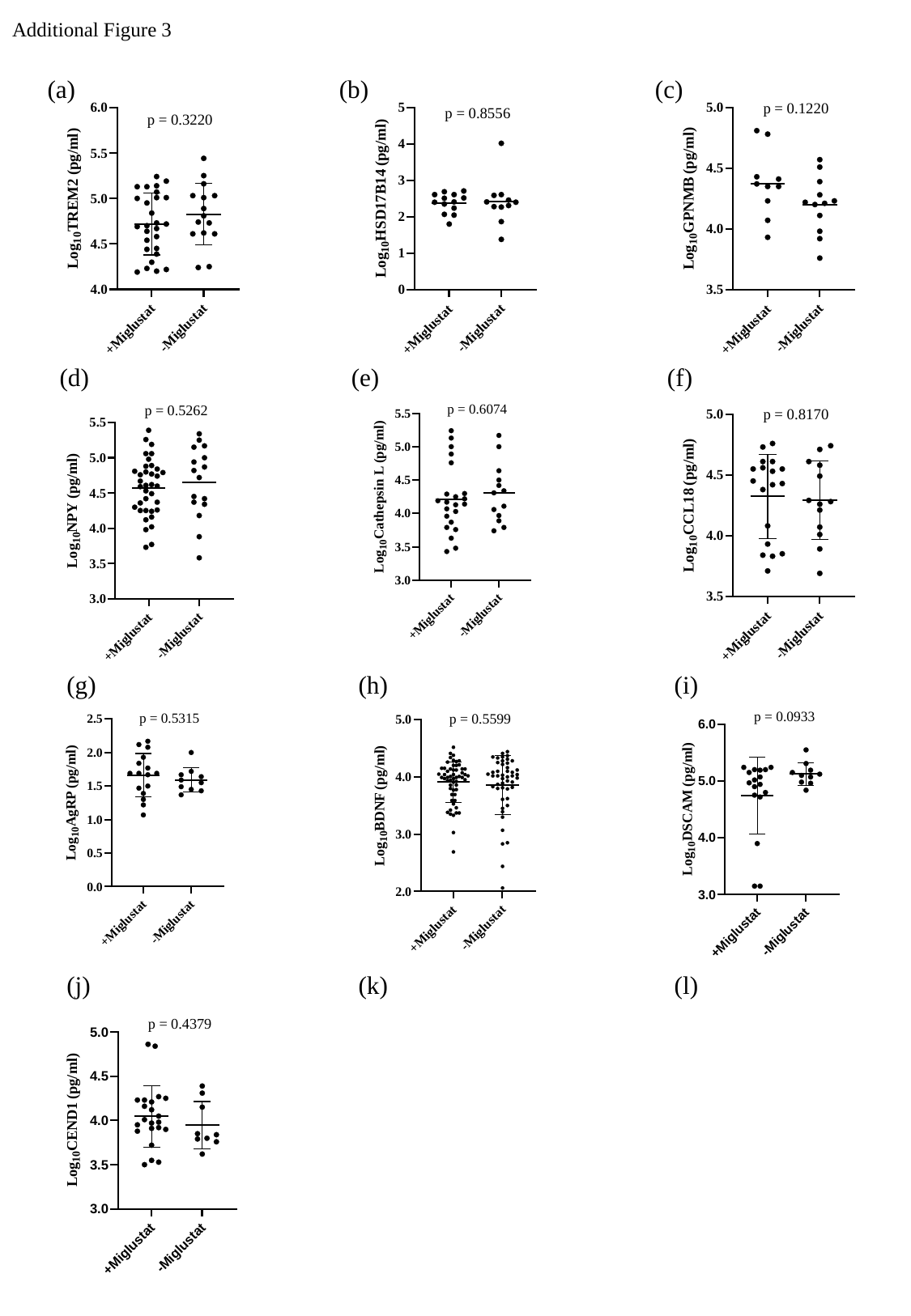

Additional Figure 3

### Slide 3
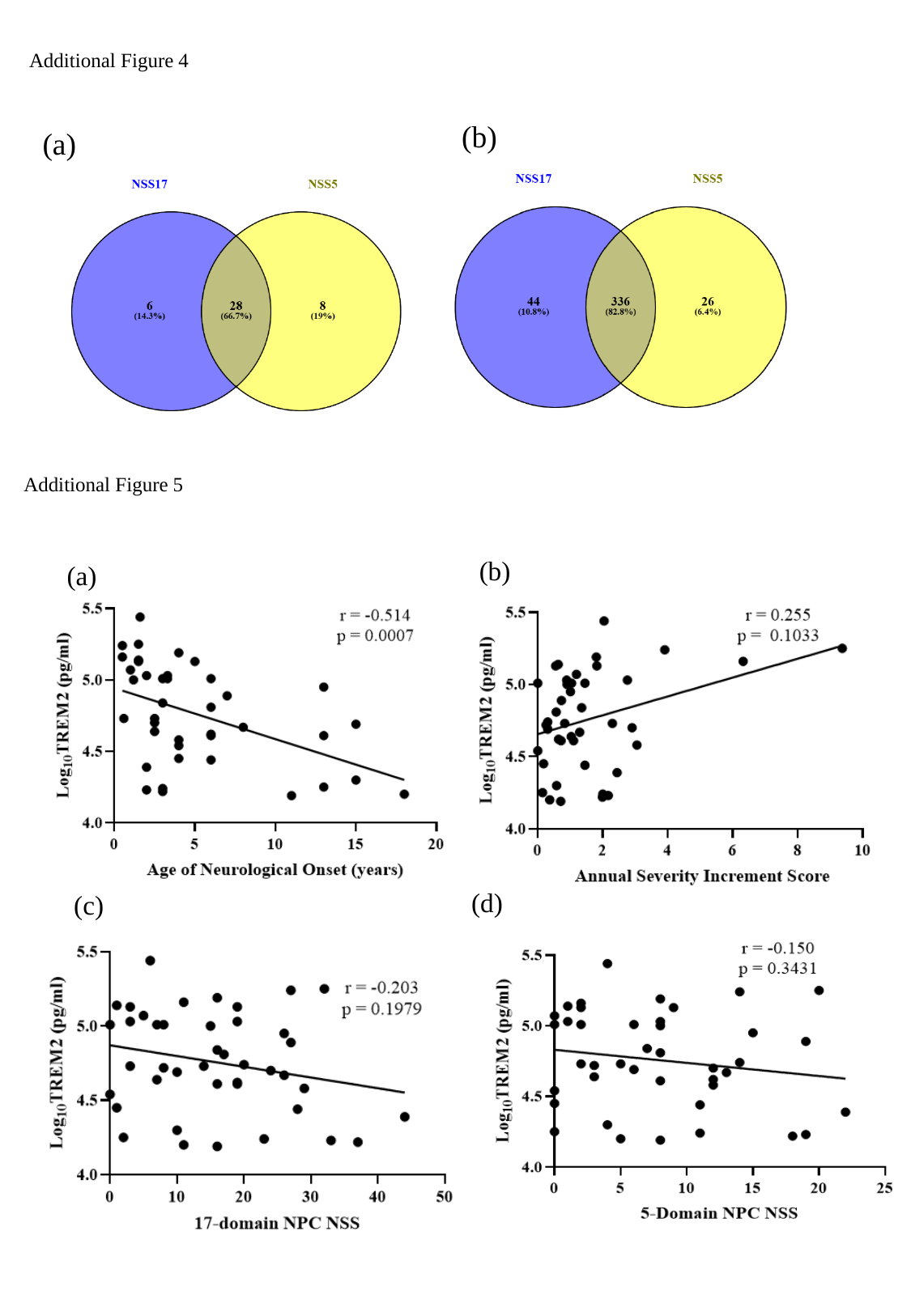

Additional Figure 4
(b)
(a)
Additional Figure 5
(b)
(a)
(d)
(c)

### Slide 4
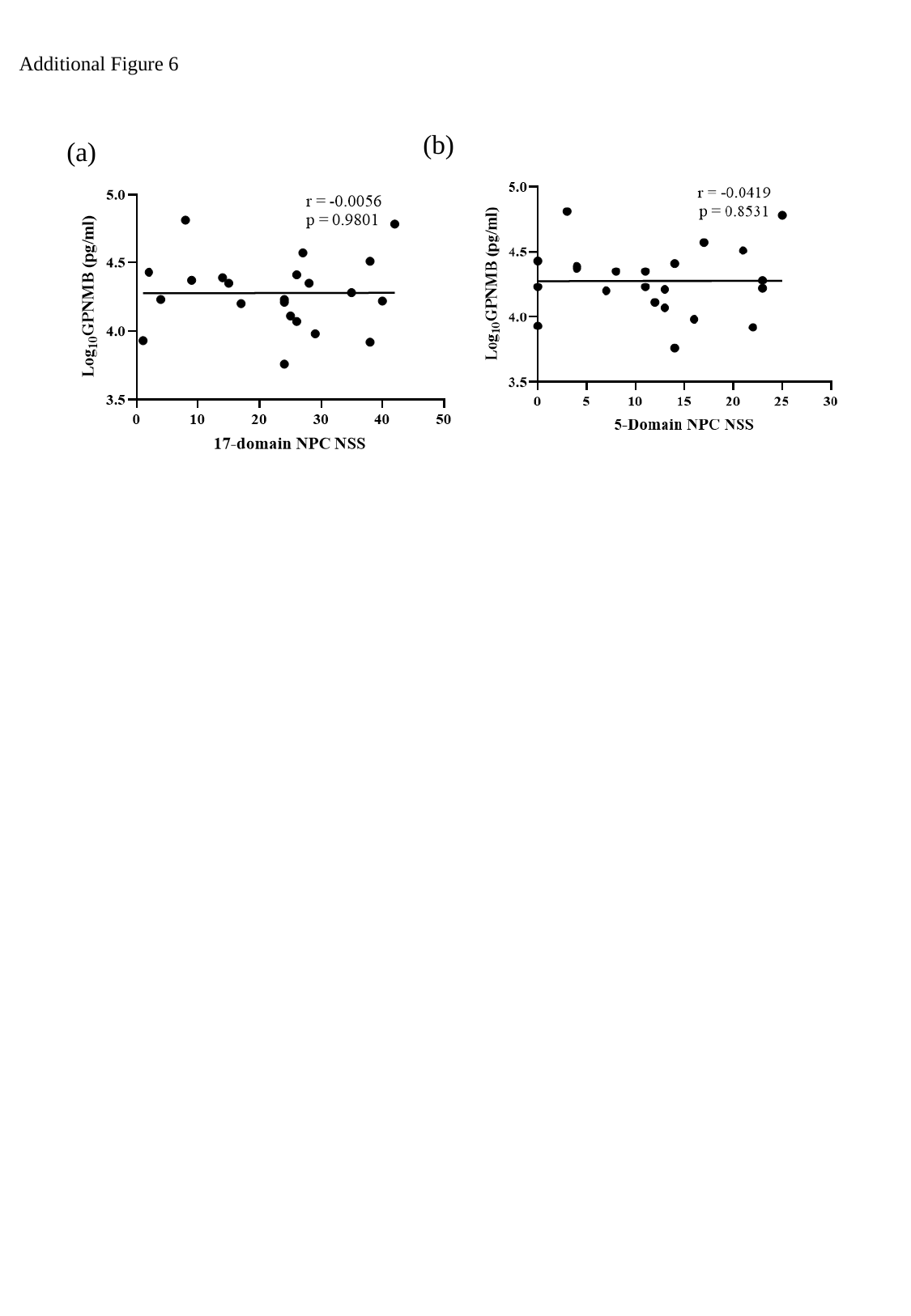

Additional Figure 6
(b)
(a)

### Slide 5
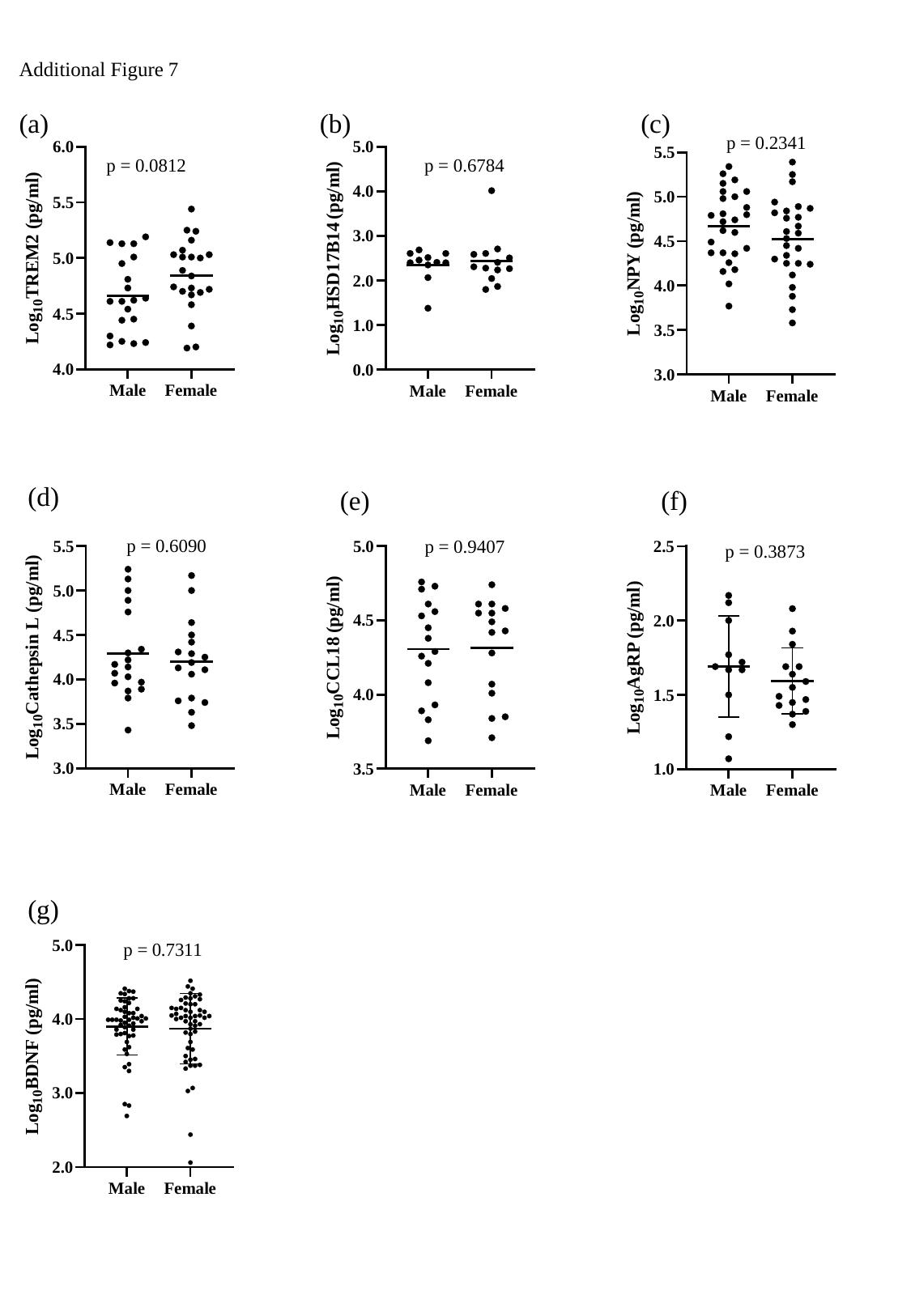

Additional Figure 7
(a)
(b)
(c)
(d)
(e)
(f)
(g)

### Slide 6
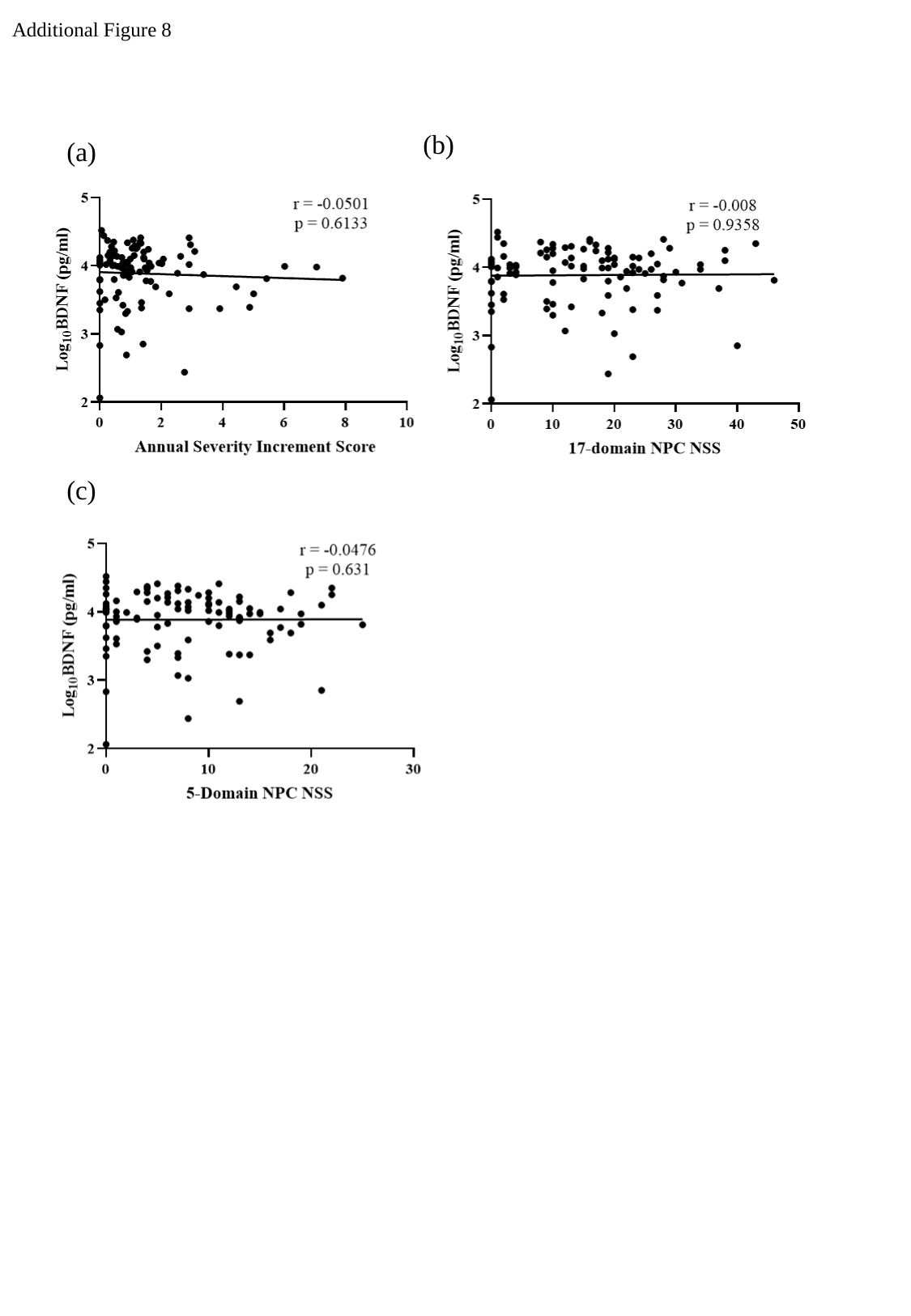

Additional Figure 8
(b)
(a)
(c)

### Slide 7
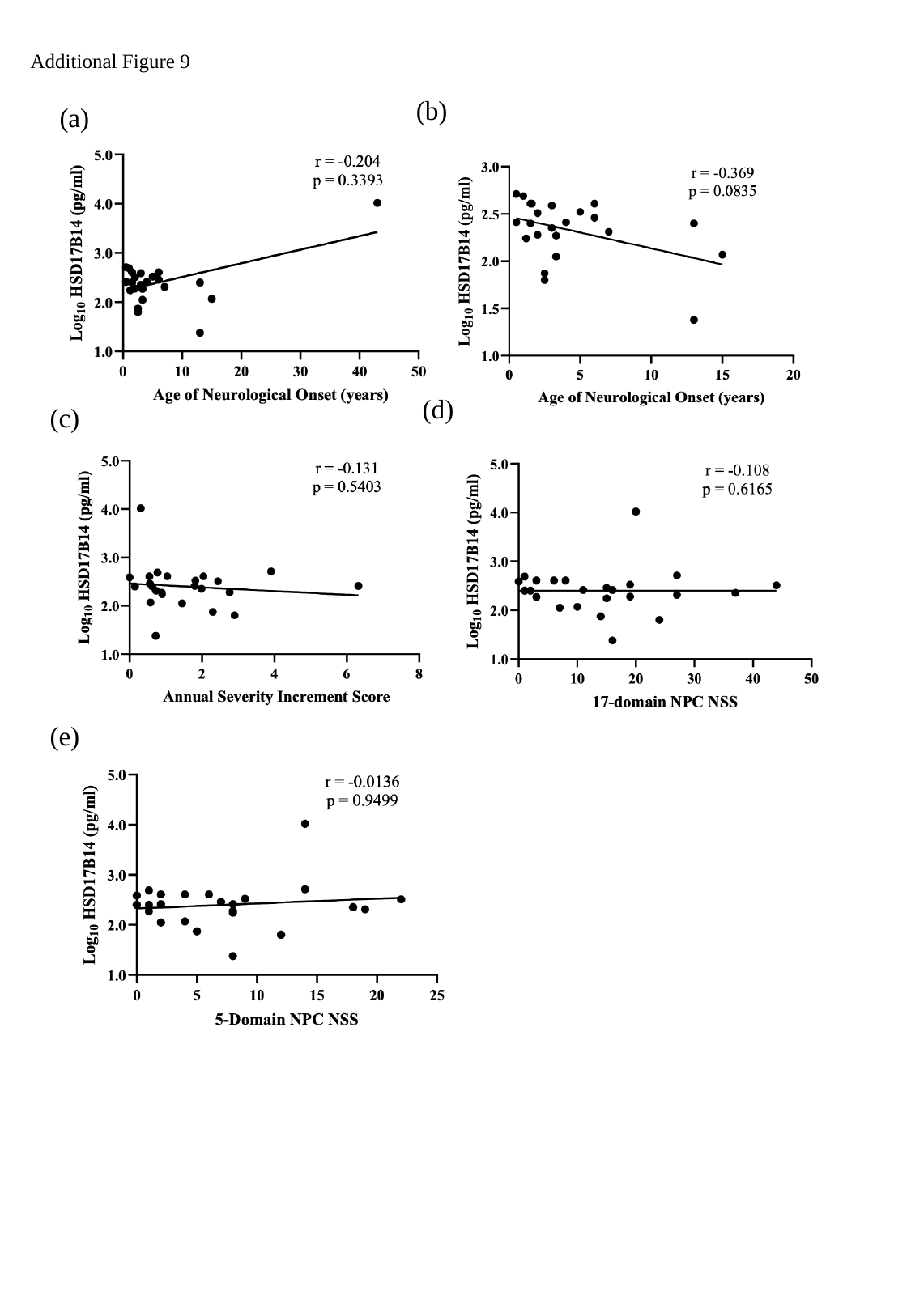

Additional Figure 9
(b)
(a)
(d)
(c)
(e)

### Slide 8
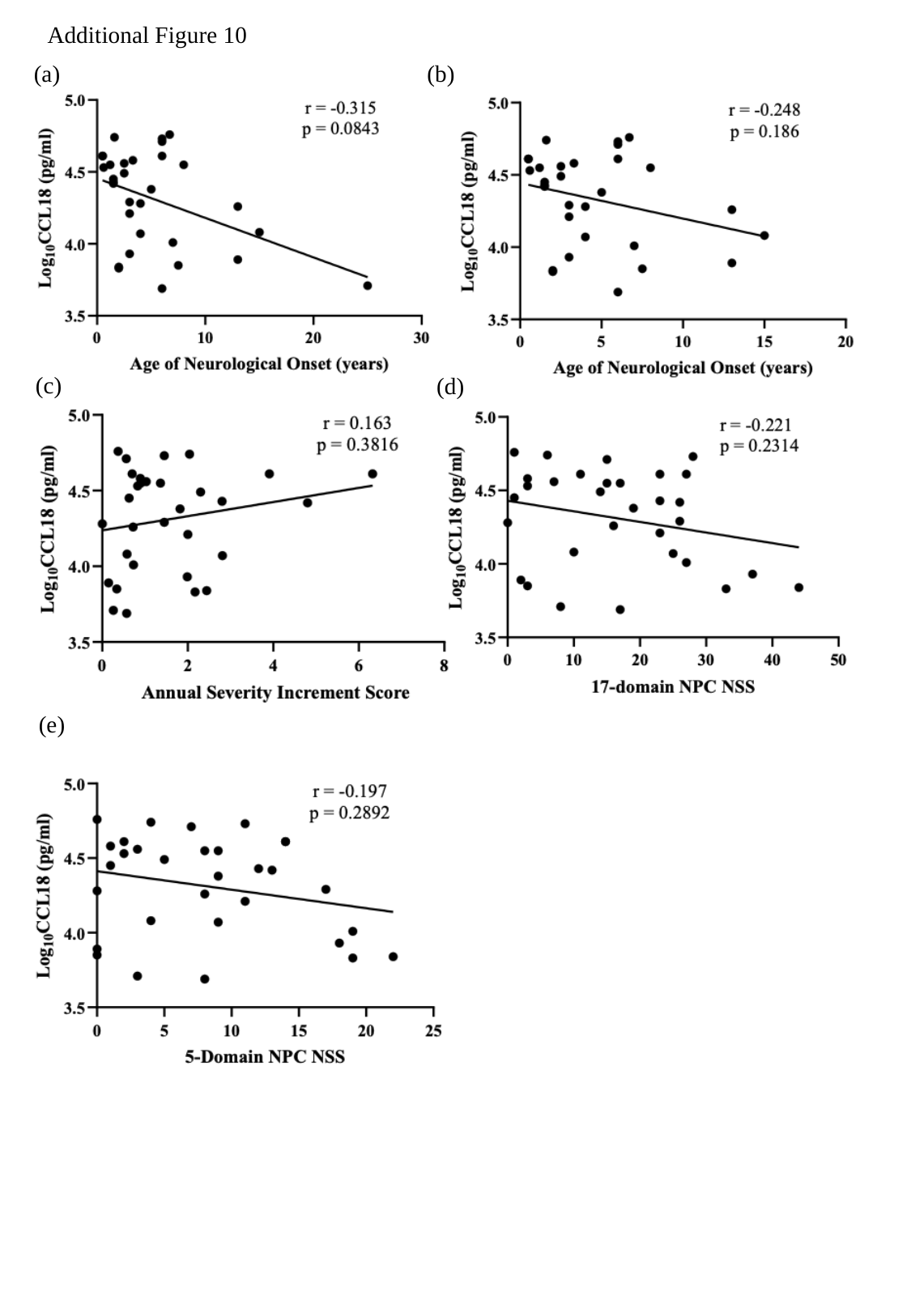

Additional Figure 10
(b)
(a)
(c)
(d)
(e)

### Slide 9
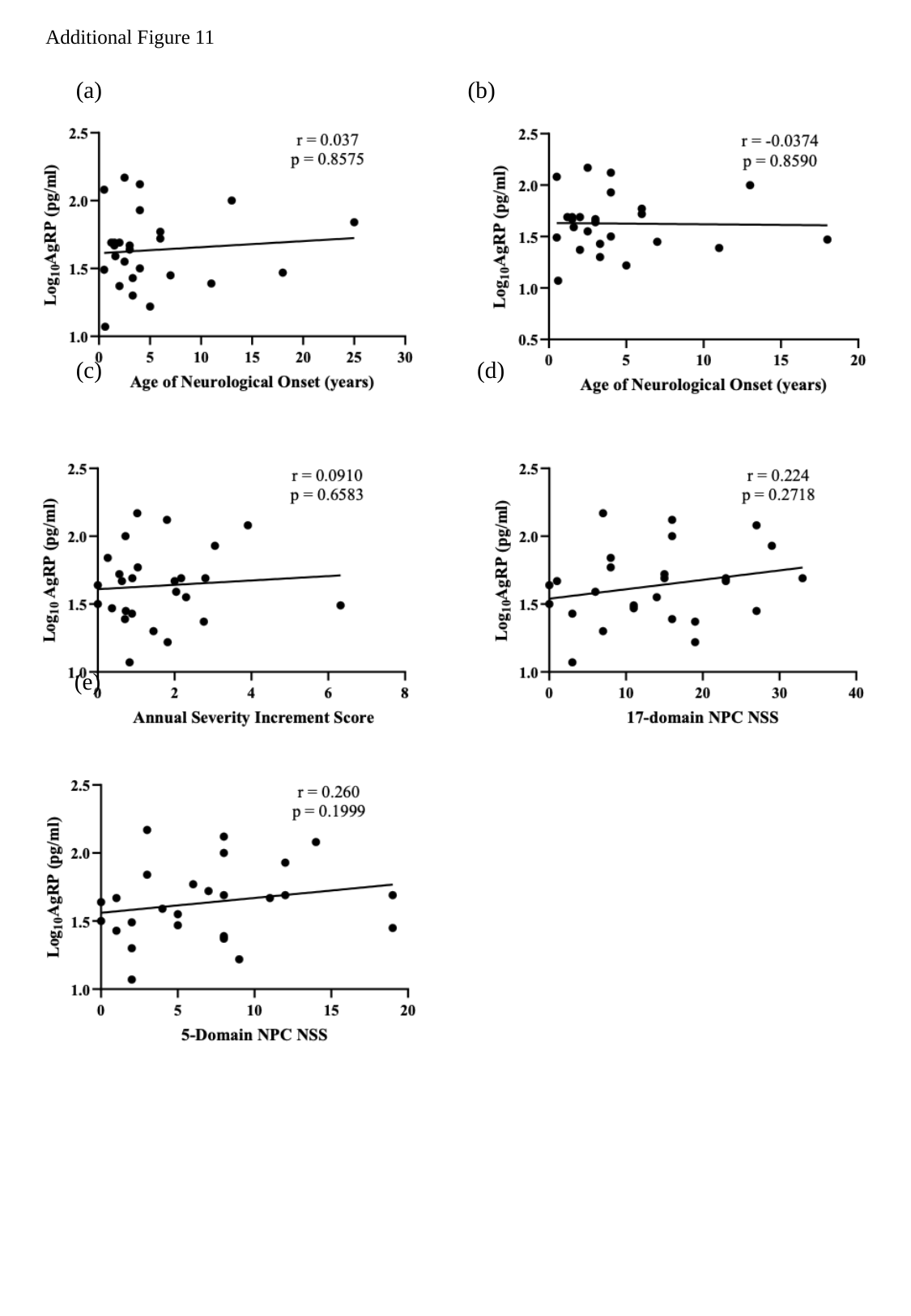

Additional Figure 11
(b)
(a)
(c)
(d)
(e)

### Slide 10
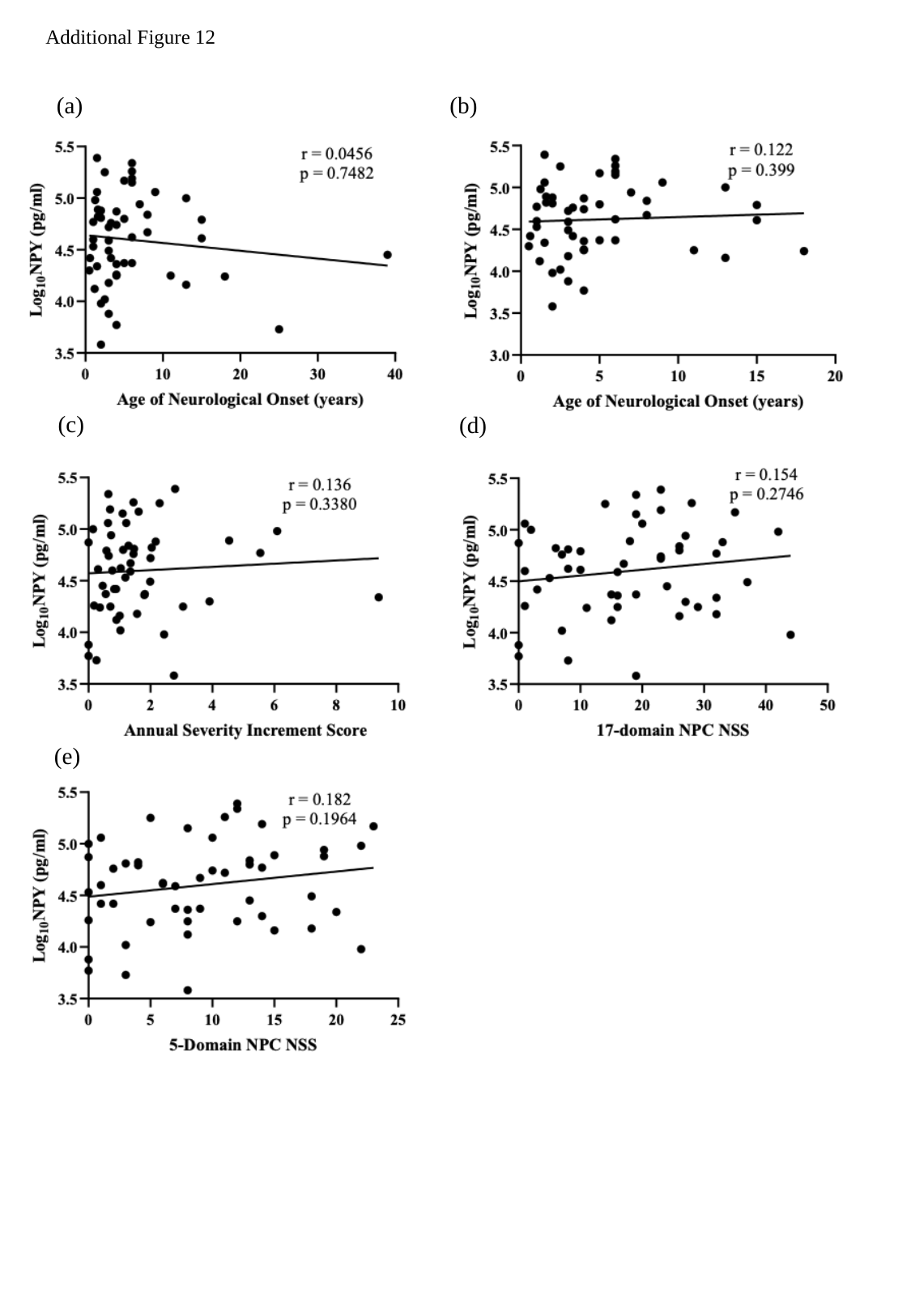

Additional Figure 12
(b)
(a)
(c)
(d)
(e)

### Slide 11
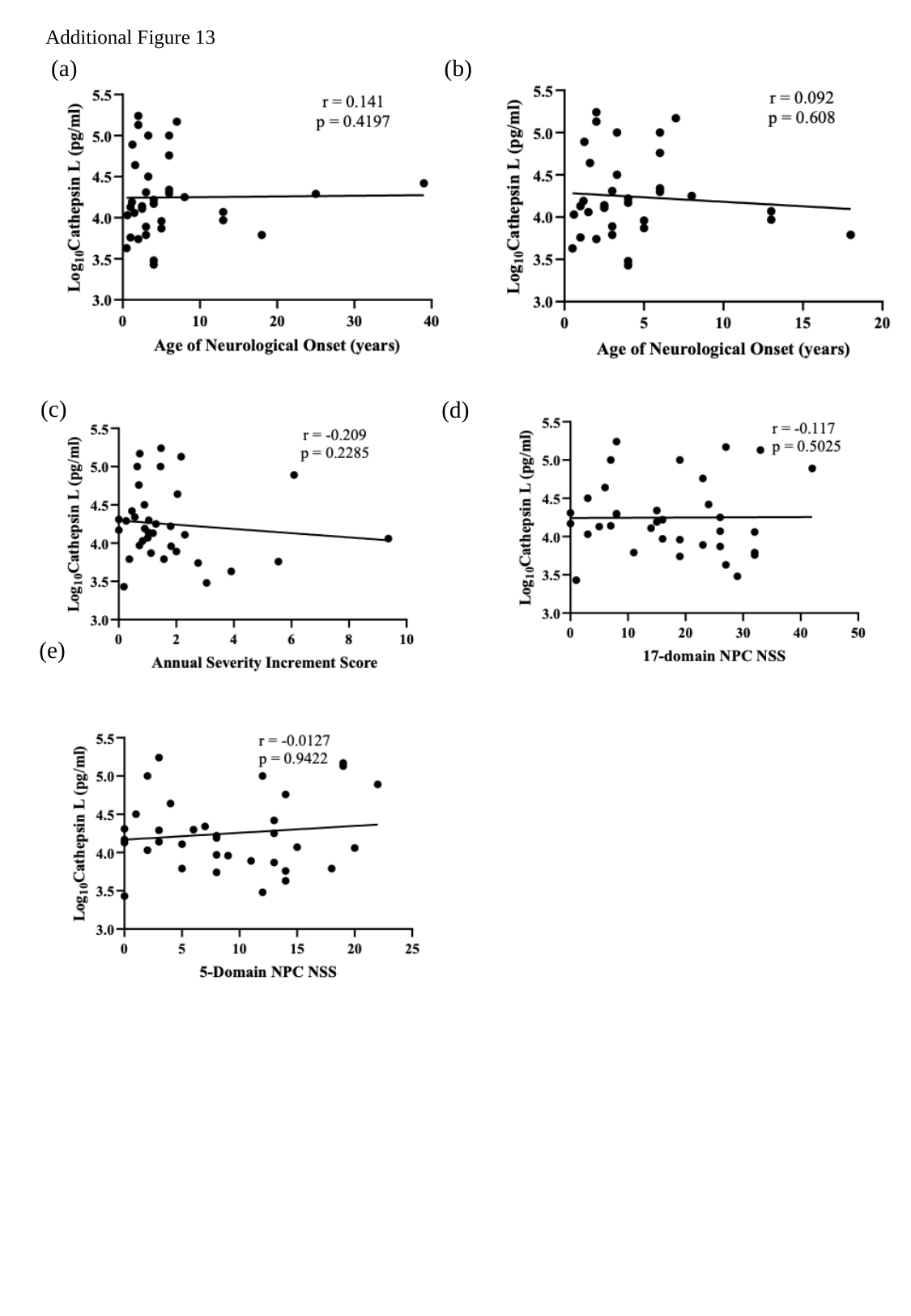

Additional Figure 13
(b)
(a)
(c)
(d)
(e)

### Slide 12
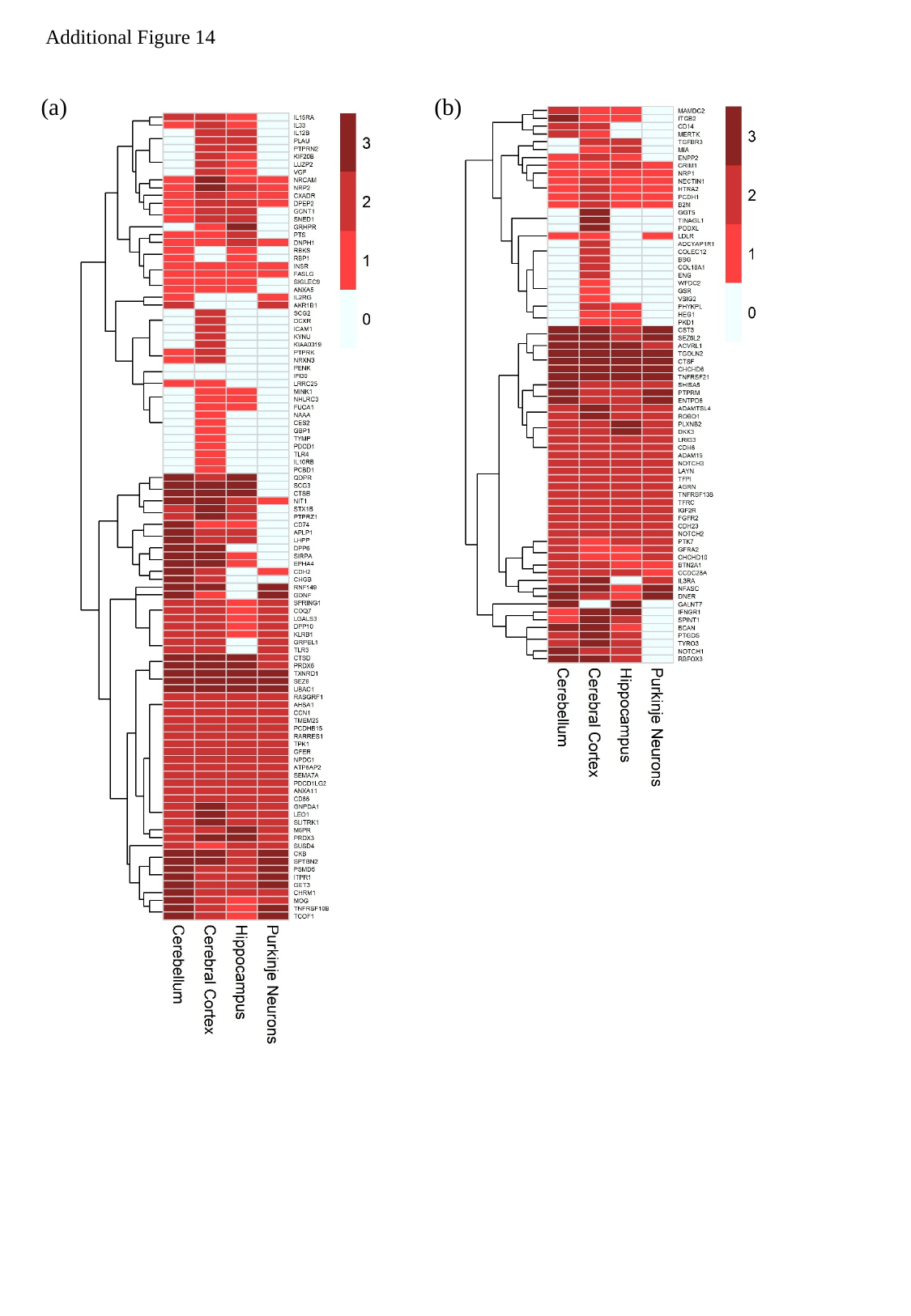

Additional Figure 14
(b)
(a)

### Slide 13
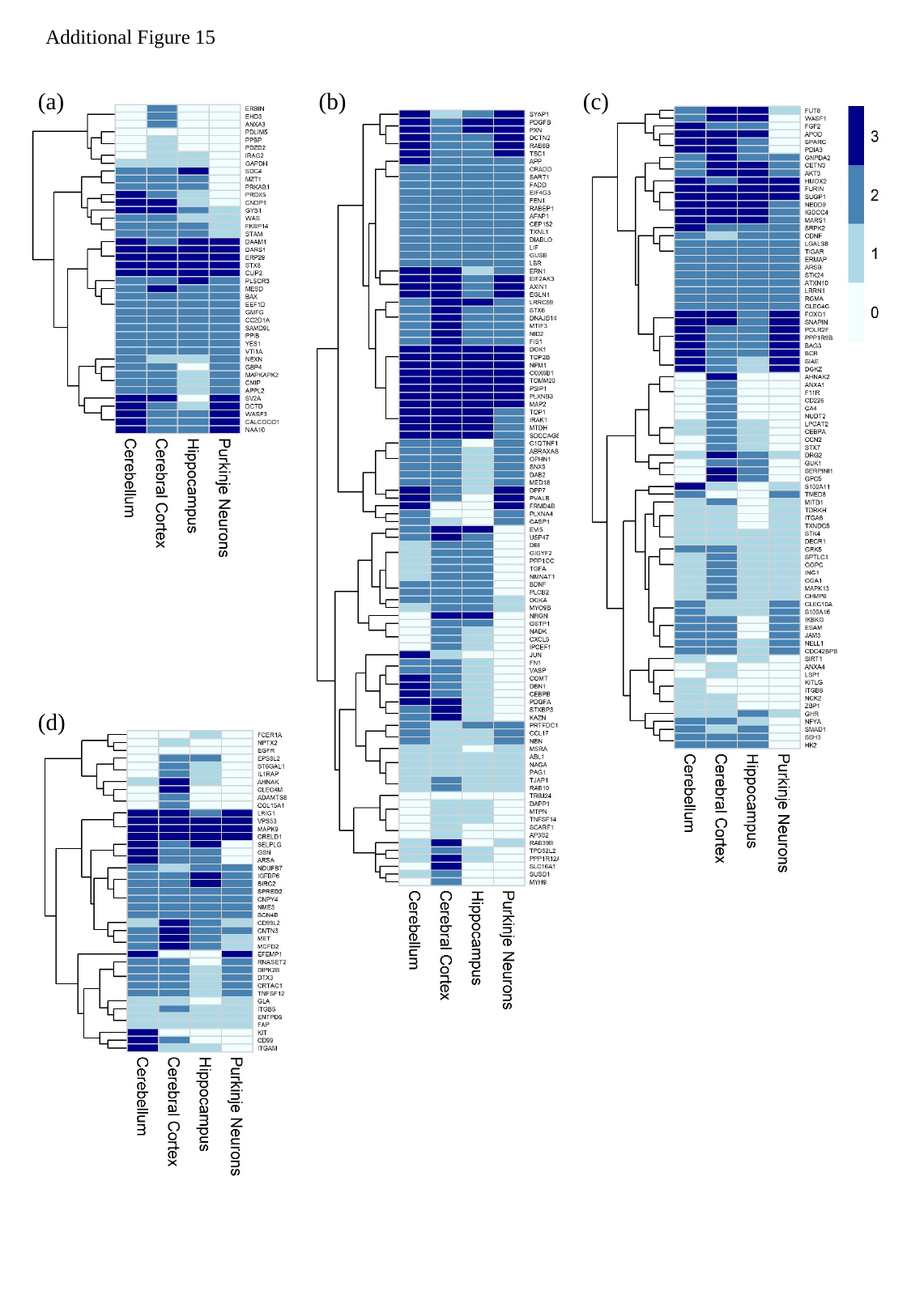

Additional Figure 15
(a)
(b)
(c)
(d)
